# Robust immune responses after one dose of BNT162b2 mRNA vaccine dose in SARS-CoV-2 experienced individuals

**DOI:** 10.1101/2021.02.07.21251311

**Authors:** Marie I. Samanovic, Amber R. Cornelius, Sophie L. Gray-Gaillard, Joseph Richard Allen, Trishala Karmacharya, Jimmy P. Wilson, Sara Wesley Hyman, Michael Tuen, Sergei B. Koralov, Mark J. Mulligan, Ramin Sedaghat Herati

## Abstract

The use of COVID-19 vaccines will play the major role in helping to end the pandemic that has killed millions worldwide. COVID-19 vaccines have resulted in robust humoral responses and protective efficacy in human trials, but efficacy trials excluded individuals with a prior diagnosis of COVID-19. As a result, little is known about how immune responses induced by mRNA vaccines differ in individuals who recovered from COVID-19. Here, we evaluated longitudinal immune responses to two-dose BNT162b2 mRNA vaccination in 15 adults who recovered from COVID-19, compared to 21 adults who did not have prior COVID-19 diagnosis. Consistent with prior studies of mRNA vaccines, we observed robust cytotoxic CD8^+^ T cell responses in both cohorts following the second dose. Furthermore, SARS-CoV-2-naive individuals had progressive increases in humoral and antigen-specific antibody-secreting cell (ASC) responses following each dose of vaccine, whereas SARS-CoV-2-experienced individuals demonstrated strong humoral and antigen-specific ASC responses to the first dose but muted responses to the second dose of the vaccine at the time points studied. Together, these data highlight the relevance of immunological history for understanding vaccine immune responses and may have significant implications for personalizing mRNA vaccination regimens used to prevent COVID-19, including booster shots.

**One Sentence Summary:** Prior history of COVID-19 affects adaptive immune responses to mRNA vaccination.

## INTRODUCTION

SARS-CoV-2 has caused hundreds of millions of infections and millions of deaths worldwide *(1)*. Although repeated infection has been described *(2, 3)*, resolution of SARS-CoV-2 infection was associated with reduced susceptibility to re-infection in animal models *(4)* and in humans *(5)*. However, it remains unknown how long this protection lasts. A number of promising vaccine candidates have emerged including mRNA vaccines, vector-based vaccines, and protein-adjuvant vaccines *(6)*. Maintenance of protective immune responses via vaccines will be important for preventing *de novo*, or recurrent, infection with SARS-CoV-2 virus.

Identification of protective correlates of immunity will be critical to predicting susceptibility to SARS-CoV-2 infection. Humoral responses have been identified as a correlate of immunity for a variety of pathogens *(7)*. In the setting of SARS-CoV-2 infection in non-human primates, humoral responses conferred protection, and T cell responses were partially protective in the setting of waning antibody titers *(8)*. Indeed, studies with mRNA vaccine candidates against SARS-CoV-2 have induced robust humoral responses against SARS-CoV-2 in animal models *(9–11)* and in humans *(12–17)* and were efficacious in large-scale clinical trials *(18, 19)*. In addition to humoral responses, mRNA vaccines induced type 1 responses in CD4^+^ T cells, as evidenced by ELISpot and intracellular cytokine staining for interferon gamma (IFNγ) and tumor necrosis factor (TNF) *(13, 14)*. Recent studies have highlighted antigen-specific B cell responses *(20)* and germinal center formation after mRNA vaccination *(21)*. However, the full spectrum of immune responses to the vaccines has not been evaluated.

Memory is the hallmark of adaptive immune responses and typically results in faster response to the pathogen upon re-exposure. Immunological history can radically shape subsequent immune responses in other ways. For example, influenza susceptibility has been linked to strain-specific exposure from decades earlier *(22, 23)*. Moreover, non-neutralizing antibody responses to acute dengue infection are a risk factor for antibody-dependent disease enhancement for serodiscordant strains *(24, 25)*. These, and other examples from the literature *(26)*, further highlight the importance for understanding immunological history in the context of COVID-19 vaccines. Moreover, large-scale clinical trials excluded individuals with a prior diagnosis of COVID-19, thereby leaving an unexplored gap in our understanding of vaccine responses in SARS-CoV-2-experienced individuals. Indeed, given the scope of the pandemic, addressing this gap in knowledge will be relevant to hundreds of millions of recovered individuals worldwide.

Here, our goal was to evaluate the effects of a prior history of COVID-19 on the immune response to mRNA vaccination. Following COVID-19, humoral and cellular immune responses persist *(27–29)*, but little is known about the effects of prior COVID-19 on subsequent exposure to SARS-CoV-2 proteins. In an observational study, we longitudinally evaluated immune responses to mRNA vaccines in adults who were naive to SARS-CoV-2 (SARS-CoV-2-naive) or who had recovered from SARS-CoV-2 (SARS-CoV-2-experienced). Using unbiased high-dimensional flow cytometry analyses, we found robust cytotoxic CD8^+^ T cell responses to vaccination but relatively muted CD4^+^ T cell responses. However, further analysis revealed subtle differences between cohorts. We found evidence for altered SARS-CoV-2-specific antibody-secreting cell (ASC) induction in circulation and altered humoral responses to vaccination depending on prior history of COVID-19. Better understanding of how prior COVID-19 shapes the immune responses to COVID-19 vaccines will improve our ability to predict susceptibility and enable personalized vaccine strategies for maintenance of immunity with booster shots.

## RESULTS

### Robust T cell responses to mRNA vaccination

Prior immune history can affect subsequent responses to antigen *(30)*. To test the effects of immunological history in the setting of COVID-19, we recruited 15 individuals who had laboratory-confirmed COVID-19 (hereafter labeled SARS-CoV-2-experienced) and 21 individuals who did not have documented COVID-19 (hereafter labeled SARS-CoV-2-naive). Participants’ ages ranged from 21 to 65, with a median age of 39 for naive adults and 43 for SARS-CoV-2-experienced individuals (table S1). All SARS-CoV-2-experienced adults had mild COVID-19 or asymptomatic infection except one individual who had severe disease (table S2). Two individuals were infected with SARS-CoV-2 within 30 days prior to vaccination, whereas the remaining 13 were at least six months beyond diagnosis of COVID-19. For these two cohorts, all participants received two doses of the BNT162b2 mRNA vaccine in accordance with the FDA Emergency Use Authorization. Peripheral immune responses were assessed before and after each dose of vaccine (**Fig. 1A**). Relative to the start of vaccination, samples were categorized as Baseline, Post 1st dose (6–13 days after vaccination), Pre 2nd dose (immediately prior to second vaccination and ∼21 days since initial vaccination), Post 2nd dose (7–12 days after second vaccination), and One month post 2nd dose (∼4 weeks after second vaccination) (**Fig. 1A** and fig. S1A). We opted to look at one week after second vaccination as that was the peak of the humoral response following mRNA vaccination *(12, 16, 31)*.

**Fig. 1.**
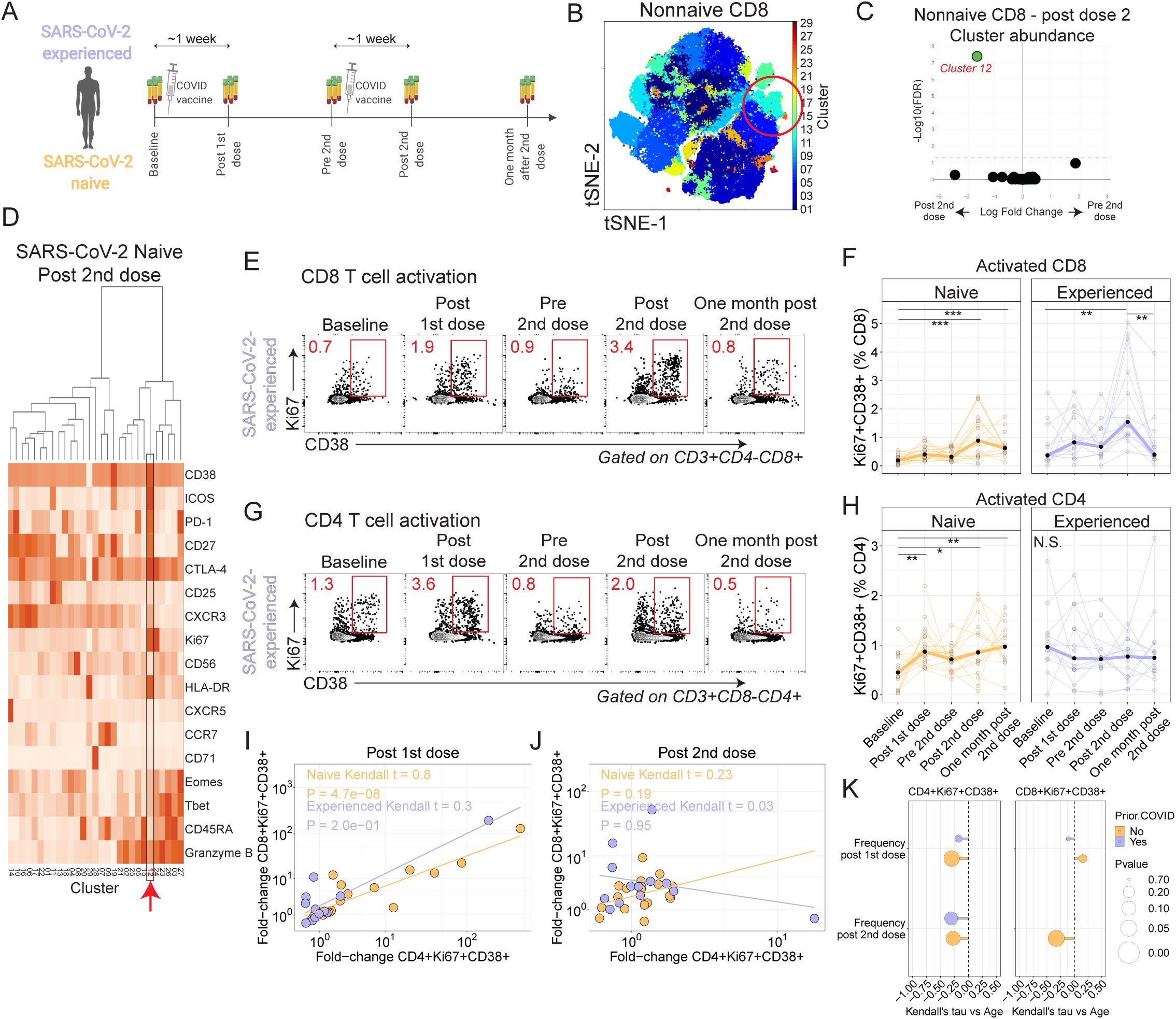
mRNA vaccination induces CD4^+^ and CD8^+^ T cell responses. (**A**) Study schematic. (**B**) Non-naive CD8^+^ T cells from all participants were colored using Phenograph clusters and projected using tSNE. Circled region indicates cluster 12. (**C**) Phenograph cluster abundance for non-naive CD8^+^ T cells was compared using edgeR for all participants before and after the second vaccination. (**D**) Heatmap for non-naive CD8^+^ T cell cluster protein expression for SARS-CoV-2-naive participants after the second vaccination. (**E**) Example of CD8^+^ T cell expression of Ki67 and CD38 in a naive participant. (**F**) Summary data for Ki67^+^CD38^+^ expression in CD8^+^ T cells by cohort. \**P* < 0.05, \*\**P* < 0.01, and \*\*\**P* < 0.001. (**G**) Example of CD4+ T cell expression of Ki67 and CD38 in a naive participant. Red number indicates frequency. (**H**) Summary data for Ki67^+^CD38^+^ expression in CD4^+^ T cells by cohort. \*\**P* < 0.01 and \*\*\**P* < 0.001. (**I and J**) Kendall rank correlations shown for the fold-changes were calculated for CD8^+^Ki67^+^CD38^+^ and CD4^+^Ki67^+^CD38^+^ T cells at one week after first dose compared to baseline (I) or at one week after second dose compared to Pre 2nd dose time point (J). (**K**) Kendall correlation for the comparison of CD4^+^Ki67^+^CD38^+^ subset versus age. Nominal *P*-values shown.

To determine the phenotype of circulating T cells responding to vaccination, we performed high-dimensional spectral flow cytometry longitudinally for all participants (fig. S1B and table S3). We initially reasoned that T cell responses would be most evident following the second dose, thus we performed cluster analysis *(32)* and tSNE representation of all non-naive CD8^+^ T cells pooled from all time points and all participants (**Fig. 1B**). Of the 29 clusters identified, only a single cluster (Cluster 12) increased in abundance at the Post 2nd dose time point compared to Pre 2nd dose (**Fig. 1C** and fig. S1C). Cells in Cluster 12 expressed high levels of multiple proteins associated with activation, including Ki67, CD38, and ICOS (**Fig. 1D** and fig. S1D). We next assessed these cells longitudinally using manual gating for Ki67 and CD38. Indeed, we found that vaccination was associated with robust induction of Ki67^+^CD38^+^ CD8^+^ T cells one week after each vaccination (**Fig. 1, E** and **F**), which was consistent with prior reports of robust induction of cytotoxic T cells after vaccination *(33)*. Compared to baseline Ki67^+^CD38^+^ CD8^+^ T cell frequencies, the first vaccination induced a median 1.9-fold increase for SARS-CoV-2-naive and 1.6-fold increase for SARS-CoV-2-experienced individuals one week post first dose. However, compared with the Pre 2nd dose time point, the second vaccination induced a 2.2-fold and 3.3-fold increase in SARS-CoV-2-naive and -experienced subjects, respectively, at one week post second dose. We also considered whether there might be differential timing of CD8^+^ T cell responses between the two cohorts, but analysis of time as a continuous variable did not identify a consistent pattern (fig. S1, E and F). Moreover, Ki67^+^CD38^+^ CD8^+^ T cells expressed high levels of Granzyme B, suggesting strong cytotoxic potential, and responded with memory kinetics to repeat exposure to SARS-CoV-2 antigens (fig. S1, G and H). Together, these data show that mRNA vaccination was associated with cytotoxic CD8^+^ T cell responses in both cohorts.

We next asked if similar changes were evident in circulating CD4^+^ T cells. Here, cluster analysis of non-naive CD4^+^ T cells identified 22 clusters, two of which increased in abundance after the second dose of vaccine: Clusters 13 and 21 (fig. S1, I and J). Of these responding clusters, Cluster 13 was associated with high expression of Ki67, CD38, and ICOS (fig. S1, K and L). Indeed, longitudinal analysis revealed induction of Ki67^+^CD38^+^ CD4^+^ T cells following immunization in the SARS-CoV-2-naive adults, with a 1.9-fold increase after first vaccination compared to baseline and a 1.3-fold increase at Post 2nd dose compared to Pre 2nd dose time points (**Fig. 1, G and H**). In contrast, we observed muted CD4^+^ responses in SARS-CoV-2-experienced adults. We considered whether there might be differential timing of CD4^+^ T cell responses between the two cohorts but again did not identify a consistent pattern (fig. S1, M and N). Interestingly, the two participants with recent COVID-19 infection show the strongest CD8^+^ T cell dynamic responses after each dose in the experienced cohort, while their CD4^+^ T cell response remains muted (fig. S1O). These data demonstrated induction of an activated CD4^+^ T cell population after vaccination in SARS-CoV-2-naive adults with only muted responses in SARS-CoV-2-experienced adults.

We next asked if these activated CD4^+^ and CD8^+^ T cell responses were correlated. Indeed, we found strong positive correlation between activated CD4^+^ and CD8^+^ responses after the first dose of vaccine and a weak correlation after the second dose of vaccine in SARS-CoV-2-naive adults (**Fig. 1, I and J**). In contrast, activated CD4^+^ and CD8^+^ responses in SARS-CoV-2-experienced adults had only a modest correlation after the first dose and no correlation after the second dose of vaccine. We also considered other demographic variables in the analysis. Aging has been associated with reduced vaccine immunogenicity and effectiveness *(34)*. Indeed, COVID-19 mortality increases with age *(35)*, and it remains unclear how well COVID-19 vaccines perform in older adults *(36)*. Here, we observed no correlation with age in activated CD8^+^ T cell responses, but for activated CD4^+^ T cell responses we found negative correlations with participant age following both primary and second vaccinations (**Fig. 1K** and fig. S1, P and Q). These results indicated the potential for reduced CD4^+^ T cell responses to vaccination with aging and underscored the need for additional studies to fully understand the effects of aging on mRNA vaccine immune responses.

### Induction of antigen-specific T cell responses after vaccination

We observed T cell responses based on phenotypic changes in both cohorts (**Fig. 1)**, which provoked the hypothesis that the vaccination had induced antigen-specific T cell responses. We first considered cytokine production following stimulation, as prior studies demonstrated the presence of TNF-or IFNγ -producing T cells following mRNA immunization *(14)*. We performed overnight stimulation of PBMC with peptide pools for the SARS-CoV-2 Spike protein across all time points, followed by intracellular staining for IFNγ or TNF. Anti-Spike T cell responses were observed at baseline (fig. S2, A to F), consistent with reports of pre-existing cross-reactivity *(37)*. Following vaccination, we observed progressively higher frequencies of CD8^+^ and CD4^+^ T cells producing these cytokines in response to peptide stimulation among SARS-CoV-2-naive adults but stable frequencies of CD8^+^ and CD4^+^ T cells producing these cytokines among SARS-CoV-2-experienced adults. These data indicated that antigen-specific T cells were induced by vaccination in SARS-CoV-2-naive adults.

We next considered other means of identifying antigen-specific cells. We *(38)* and others *(37, 39, 40)* have demonstrated the use of activation-induced markers (AIM) as a method to study antigen-specific T cells that may not produce cytokines. Following overnight stimulation of PBMC with peptide pools for the SARS-CoV-2 Spike protein, we identified induction of CD137^+^IFNγ ^+^ CD8^+^ cells after vaccination in SARS-CoV-2-naive adults but not in SARS-CoV-2-experienced adults (**Fig. 2, A and B**, and fig. S2G). We next considered CD4^+^ T cell induction by vaccination (**Fig. 2, C and D**, and fig. S2H). Similar to CD8^+^ T cell responses, CD4^+^ T cells coexpressing CD69 and CD200 after stimulation increased 9-fold at one month post second vaccination relative to baseline in SARS-CoV-2-naive adults (*P*=0.025, Kruskal-Wallis with Dunn’s post-test) but did not increase in SARS-CoV-2-experienced adults. We also considered other surface proteins including co-expression of OX40 and CD137 and observed similar results (fig. S2, I to L). In sum, Spike-specific T cell responses were induced in SARS-CoV-2-naive adults but did not further increase in SARS-CoV-2-experienced adults after mRNA vaccination.

**Fig. 2.**
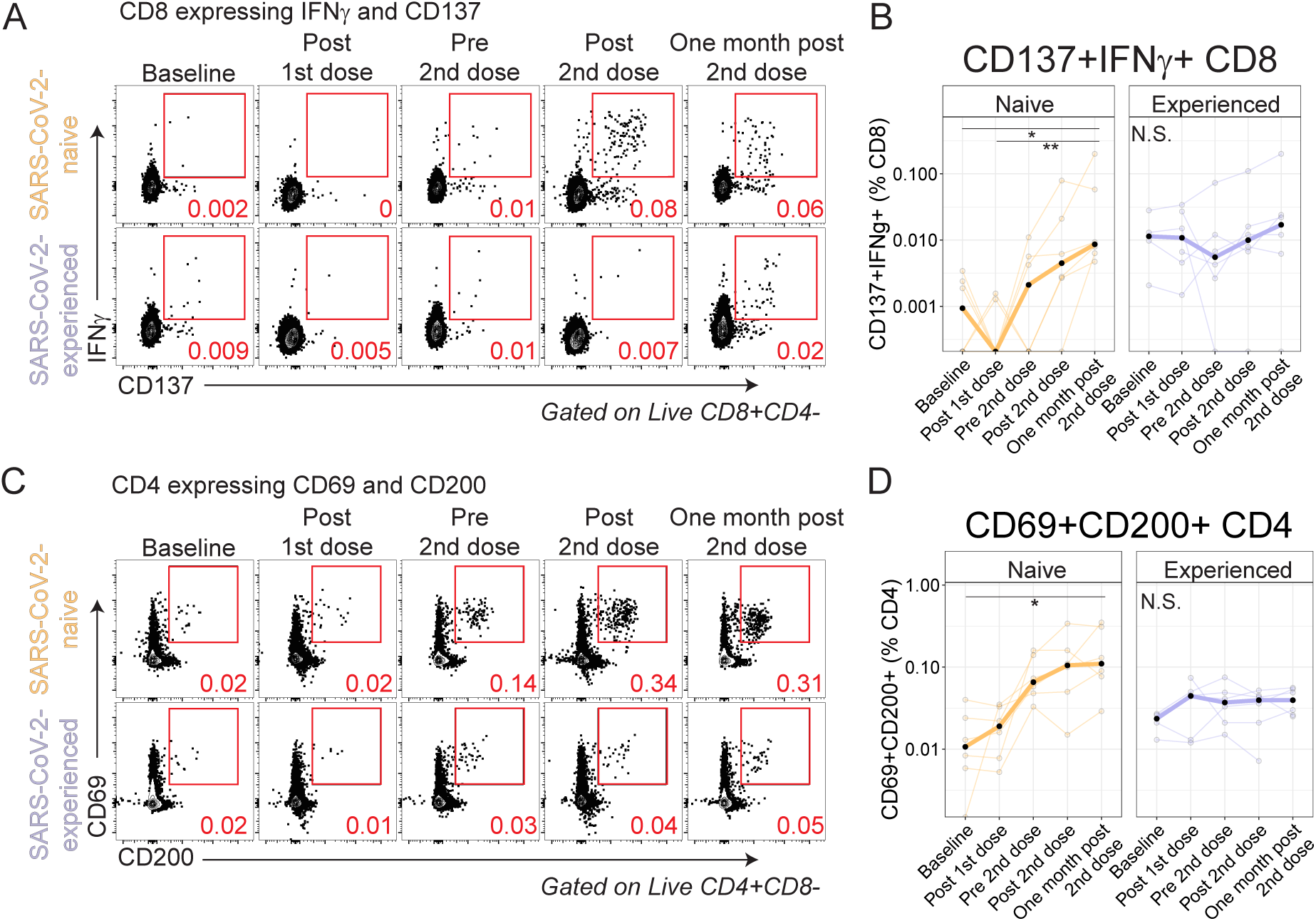
Antigen-specific T cells are induced by vaccination. PBMC were rested overnight then stimulated for 20 hours with SARS-CoV-2 Spike peptides in the presence of monensin, followed by phenotypic analysis. (**A and B**) Example CD8^+^ T cell plots for SARS-CoV-2-naive and SARS-CoV-2-experienced individuals for the expression of IFNγ and CD137 for the full time course (**A**) and summary plots (**B**). (**C and D**) Example CD4^+^ T cell plots for SARS-CoV-2-naive and SARS-CoV-2-experienced individuals for the expression of CD69 and CD200 for the full time course (C) and summary plots (D). Red number indicates frequency. \**P* < 0.05 and \*\**P* < 0.01 by Dunn’s post-test.

### Differential induction of circulating T follicular helper cells after vaccination

Most vaccines are thought to confer protection via induction of a class-switched, affinity-matured antibody response *(7)*, and, given the subtle differences in CD4^+^ T cell responses following mRNA vaccination between cohorts (**Fig. 1**), we next considered CD4 T cell responses that might be relevant to the antibody response. Maturation of B cell responses within germinal centers requires help from CD4^+^ T follicular cells (Tfh) *(41, 42)*, and indeed Spike-specific germinal center B cells were identified in axillary lymph node aspirates after mRNA vaccination *(21)*. However, lymphoid tissue is challenging to routinely study in humans. We, and others, have focused on a circulating Tfh-like subset with similar phenotypic, transcriptional, epigenetic, and functional characteristics to lymphoid Tfh *(43–47)*. Indeed, we previously found that vaccination induced antigen-specific ICOS^+^CD38^+^ circulating Tfh (cTfh) which correlated with plasmablast responses and demonstrated memory kinetics *(38)*. Furthermore, other studies identified similar activated cTfh responses in non-human primates following mRNA vaccination for influenza *(48)*. However, activated cTfh have not been evaluated in humans following SARS-CoV-2 mRNA vaccination.

We scrutinized all time points for evidence of cTfh responses. ICOS^+^CD38^+^ cTfh cells increased following vaccination in SARS-CoV-2-naive adults and peaked one week after the second vaccine dose (**Fig. 3, A and B**). In contrast, SARS-CoV-2-experienced adults did not show similar induction of cTfh cells following either dose of the vaccine (**Fig. 3B**). In prior studies, antigen-specific ICOS^+^CD38^+^ cTfh were shown to express CXCR3 following influenza vaccination *(38, 46)*, thus we next considered the subset of ICOS^+^CD38^+^ cTfh that expressed CXCR3. Here, we identified an 2.1-fold induction of CXCR3^+^ cells among ICOS^+^CD38^+^ cTfh cells in SARS-CoV-2-experienced adults after the first vaccine dose and a 2.0-fold increase among SARS-CoV-2-naive adults after the first dose (**Fig. 3, C and D**). There was minimal change in CXCR3 expression in ICOS^+^CD38^+^ cTfh one week after the second dose of vaccine in either cohort. Of note, SARS-CoV-2-experienced participants with recent COVID-19 showed a more dynamic induction of CXCR3^+^ cells among ICOS^+^CD38^+^ cTfh cells after each vaccine dose (fig. S3A).

**Fig. 3.**
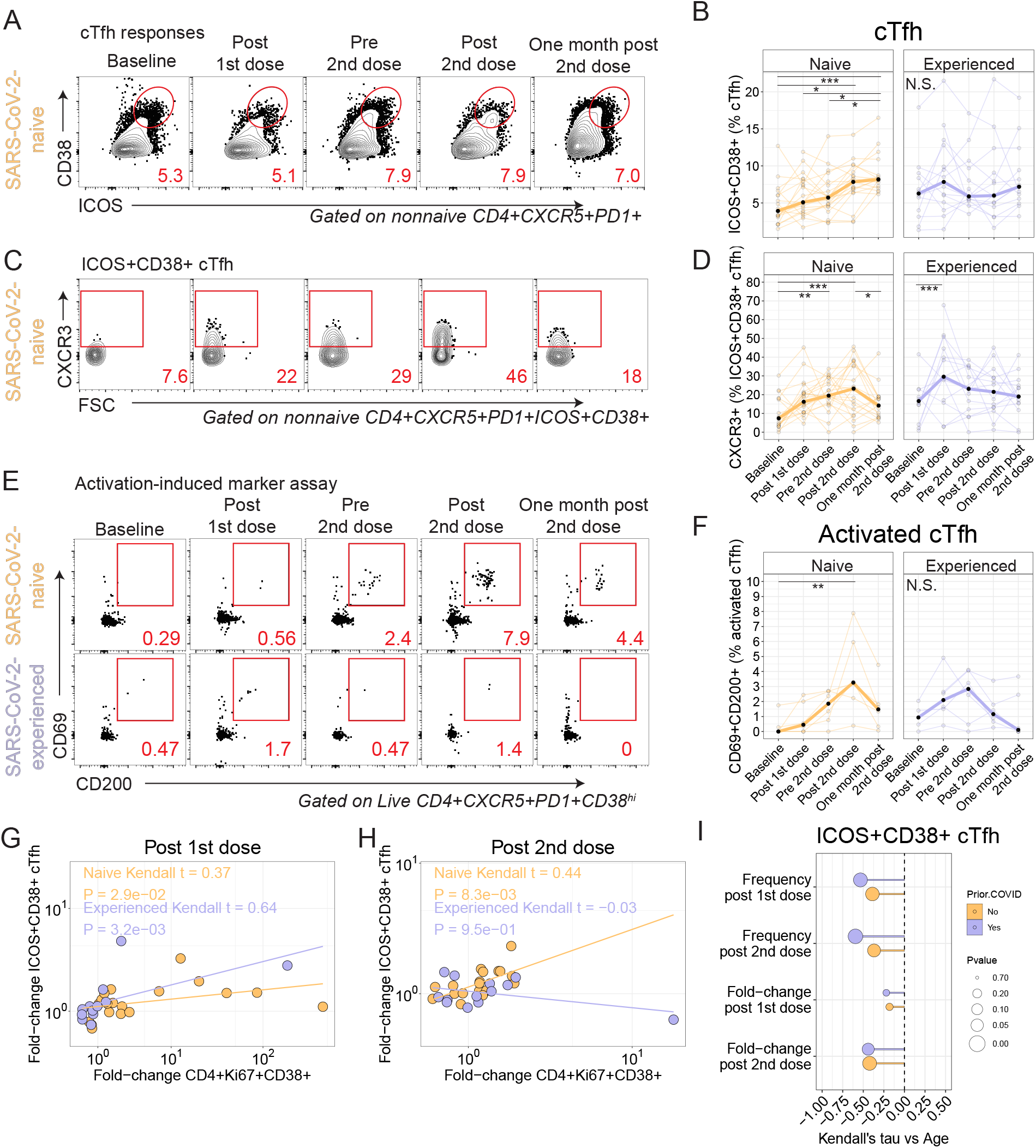
Differential induction of circulating T follicular helper cells after vaccination. For all plots, \**P* < 0.05, \*\**P* < 0.01, and \*\*\**P* < 0.001 by Dunn’s post-test unless otherwise noted. (**A**) Example naive participant shown for ICOS and CD38 in cTfh. (**B**) Summary data for expression of ICOS and CD38 in cTfh. (**C**) Example shown for CXCR3 in ICOS^+^CD38^+^ cTfh. (**D**) Summary data for CXCR3 expression in ICOS^+^CD38^+^ cTfh. (**E**) PBMC were stimulated with SARS-CoV-2 Spike peptides for 20 hours, followed by phenotypic analysis. CD38^hi^ cTfh shown for expression of CD69 and CD200. Red number indicates frequency. (**F**) Summary plot for CD69 and CD200 coexpression among CD38^hi^ cTfh. (**G and H**) Kendall correlation between the fold-change in ICOS^+^CD38^+^ cTfh and Ki67^+^CD38^+^ CD4 T cells at one week Post 1st dose compared to baseline (G) or one week after second dose compared to Pre 2nd dose (H). (**I**) Kendall correlations for the comparison of CD4^+^Ki67^+^CD38^+^ subset versus age. Fold-change Post 1st dose is relative to baseline, and fold-change Post 2nd dose is relative to Pre 2nd dose time point. Nominal *P*-values shown.

We next used the AIM assay to identify antigen-specific cTfh induced by vaccination, as we have done previously *(38)*. Spike-specific activated cTfh in the AIM assay were identified as CXCR5^+^PD1^+^ CD4 T cells that had high expression of CD38 and that coexpressed CD69 and CD200 after overnight stimulation with a Spike peptide pool (**Fig. 3, C to F**). In this subset, we found a 14.5-fold induction of cells that among SARS-CoV-2-naive adults at the Post 2nd dose time point relative to baseline (*P*=0.008, Kruskal-Wallis with Dunn’s post-test), whereas no increase was observed among SARS-CoV-2-experienced adults during the same time interval (**Fig. 3, E and F**), which was similar to the trend observed for the CXCR3^+^ICOS^+^CD38^+^ cTfh in **Fig. 3, C and D**. Together, these data demonstrated the induction of Spike-specific cTfh in SARS-CoV-2-naive adults following mRNA vaccination.

Thus, we asked if the cTfh response correlated with the Ki67^+^CD38^+^ CD4^+^ response. Indeed, ICOS^+^CD38^+^ cTfh from SARS-CoV-2-naive adults correlated positively with Ki67^+^CD38^+^ CD4^+^ T cells for the fold-change at Post 1st dose compared to baseline (**Fig. 3G** and fig. S3B) and at Post 2nd dose compared to Pre 2nd dose (**Fig. 3H** and fig. S3C). In contrast, SARS-CoV-2-experienced adults had a positive correlation after the first dose and did not have a correlation after the second dose. We also found negative correlations with age in both cohorts (**Fig. 3I**), similar to what was observed for activated CD4^+^ responses (**Fig. 1**).

We also evaluated other well-established cellular correlates of the humoral response such as plasmablasts *(48)*, CD21^lo^ B cells *(49)*, and CD71^+^ B cells *(50)* but found little or no induction of these subsets in either cohort longitudinally (fig. S3, D to J). Plasma CXCL13, which has been reported as a plasma biomarker of early germinal center activity *(51)*, also did not change following vaccination in either cohort (fig. S3, K and L).

Altogether, we found antigen-specific induction of ICOS^+^CD38^+^ cTfh following vaccination with subtle differences between cohorts. Indeed, while the ICOS^+^CD38^+^ cTfh frequency continued to increase in the SARS-CoV-2-naive adults there was no evidence of sustained induction of cTfh in SARS-CoV-2-experienced adults over the course of the vaccination series. Given that Tfh provide help to B cells, these data provoked the question as to whether B cell responses also differed by prior history of COVID-19.

### Induction of SARS-CoV-2-specific ASC in circulation after vaccination

We observed subtle differences in induction of ICOS^+^CD38^+^ cTfh following vaccination based on prior history of COVID-19 (**Fig. 3**). Thus, we next asked if antigen-specific B cell responses induced by vaccination were influenced by prior exposure to the virus. To test this, we performed enzyme-linked immunospot (ELISpot) analyses of antibody-secreting cells (ASC) for reactivity against SARS-CoV-2 proteins one week after each vaccine dose for both cohorts.

Given the persistence of SARS-CoV-2-reactive B cells in individuals who recovered from COVID-19 *(28)*, we expected to find a stronger antigen-specific ASC response in SARS-CoV-2-experienced adults than SARS-CoV-2-naive adults after the first dose of vaccine. Indeed, after the first dose of vaccine, SARS-CoV-2-naive adults had few SARS-CoV-2-specific ASCs detected, whereas SARS-CoV-2-experienced adults had stronger IgG-secreting ASC responses to RBD, S1, and S2 proteins (**Fig. 4, A** to **C**, fig. S4, A and B). Moreover, IgA-secreting ASC were identified predominantly in SARS-CoV-2-experienced adults after the first vaccine dose, albeit at a somewhat lower frequency than IgG-secreting ASC (**Fig. 4D**). Few IgM-secreting ASC were identified (fig. S4C). Although global plasmablast frequencies did not change with vaccination (fig. S3, D to F), we did indeed find evidence of antigen-specific ASC responses following the first vaccine dose among both cohorts.

**Fig. 4.**
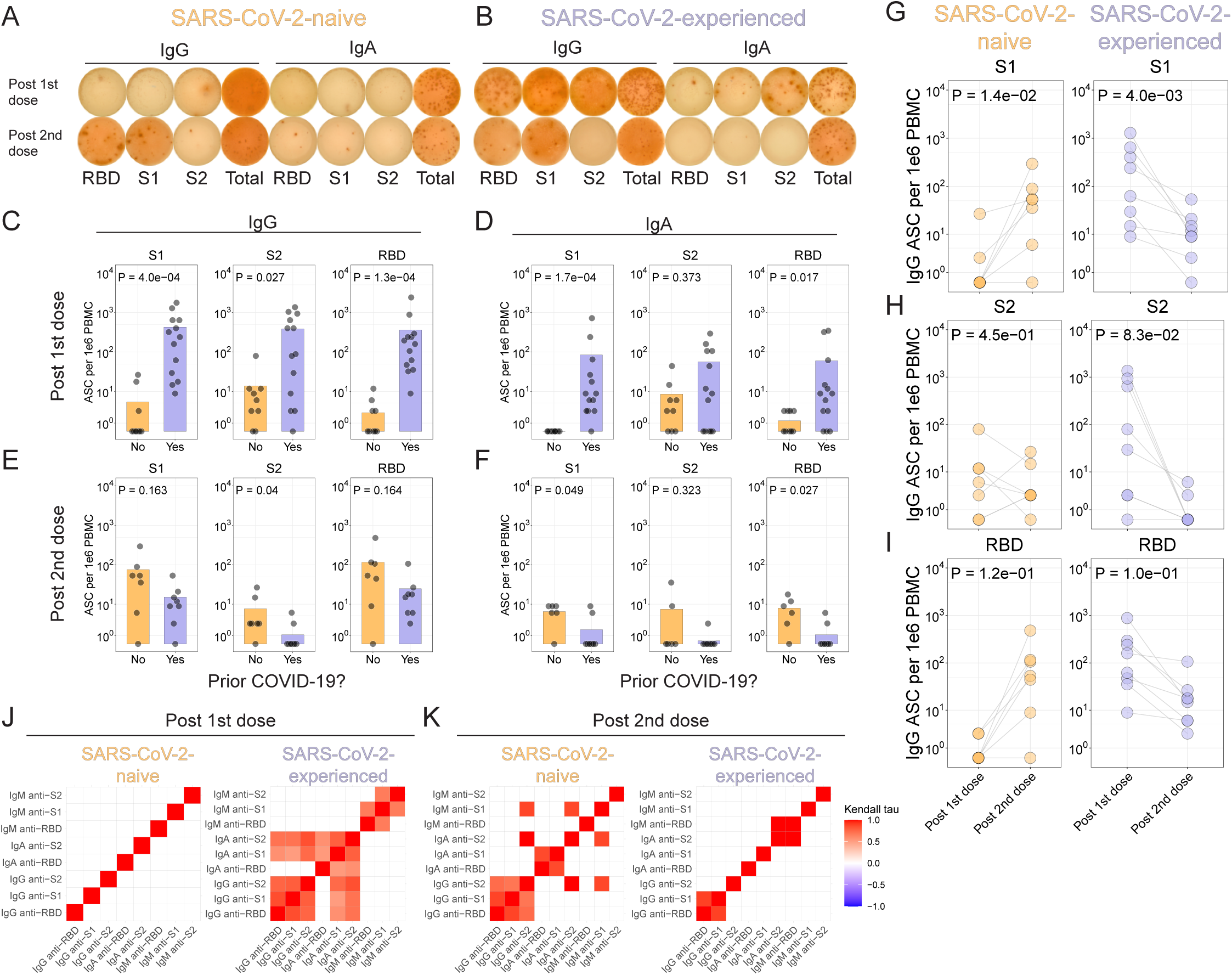
Few antigen-specific ASC induced in circulation after the second vaccine dose in SARS-CoV-2-experienced adults. (**A and B**). Antibody-secreting cell (ASC) ELISpots for a SARS-CoV-2-naive (A) or SARS-CoV-2-experienced (B) adult one week after each dose of vaccine. (**C to F**) Summary statistics for ELISpot assays. For each panel, S1 (left), S2 (middle), or RBD (right) antigens for IgG or IgA are represented, at one week after first dose (C and D) or second dose (E and F). Nominal *P* values from Wilcoxon tests. (**G to I**) ELISpot results for SARS-CoV-2-naive (left) or SARS-CoV-2-experienced (right) adults for S1 (**G**), S2 (**H**), or RBD (**I**). Connected lines indicate repeated measurements from the same participants. Nominal *P* values from paired t-tests. (**J and K**) Kendall correlations for ELISpot results one week after the first vaccination (J) or one week after the second vaccination (K). Correlations shown for comparisons with nominal *P* values <0.05.

We next asked if the second vaccination also induced strong antigen-specific ASC responses in the two cohorts. Indeed, the second dose of vaccine robustly induced S1-and RBD-reactive ASC in SARS-CoV-2-naive adults (**Fig. 4, E to I**). In contrast, however, the second dose of vaccine induced similar, or weaker, ASC responses in SARS-COV-2-experienced adults approximately one week after vaccination for all three SARS-CoV-2 antigens tested (fig. S4D). Spike-specific ASC induction was correlated by isotype and antigen in SARS-CoV-2-experienced adults one week after the first vaccination and SARS-CoV-2-naive one week after the second vaccination (**Fig. 4, J and K** fig. S4, E and H). However, correlations by isotype and antigen were not observed in the SARS-CoV-2-experienced adults following the second vaccination. Furthermore, the pattern of these correlations changed between doses. For example, among SARS-CoV-2-naive adults, there was correlation between IgM-, IgA-, and IgG-secreting ASC after the second vaccination but not after first vaccination, whereas similar correlations were observed in ASC from SARS-CoV-2-experienced adults after the first, but not after the second, vaccination. The lack of correlation may be partly due to the low numbers of ASC detected, but further studies will be needed to determine how unswitched and switched B cell populations respond in the settings of priming and recall responses.

Together, these data demonstrated increased induction of antigen-specific ASC responses with repeated vaccination in SARS-CoV-2-naive adults, whereas fewer antigen-specific ASC were observed in circulation with repeated vaccination in SARS-CoV-2-experienced adults.

### Antigen-specific B cells are induced by vaccination

To further study the induction of SARS-CoV-2-specific B cell responses after immunization, we used fluorescent recombinant RBD protein to identify RBD-reactive B cells in PBMC (fig. S5A). We found RBD-reactive B cells were 2-fold higher at baseline among SARS-CoV-2-experienced adults compared to SARS-CoV-2-naive adults (*P*=0.03, Wilcoxon test) (fig. S5B). Furthermore, fewer RBD-reactive B cells were class-switched B cells in SARS-CoV-2-naive adults, as evidenced by expression of IgG, relative to SARS-CoV-2-experienced adults (*P*=0.02, Wilcoxon test) (fig. S5C). Following immunization, increased frequencies of RBD^+^ B cells were observed in both cohorts (**Fig. 5, A and B**), with fold-changes of 2.9 and 4.8 in SARS-CoV-2-naive and SARS-CoV-2-experienced adults, respectively, at Post 2nd dose relative to Baseline. Moreover, SARS-CoV-2-naive adults had progressively more IgG^+^ RBD^+^ B cells at Post 2nd dose than at Baseline (*P*=0.01, Kruskal-Wallis with Dunn’s post-test) (**Fig. 5, C and D**), whereas SARS-CoV-2-experienced adults had minimal change in the proportion of IgG^+^ RBD^+^ B cells at the same time points (*P*=0.24, one-way ANOVA with Tukey’s post-test).

**Fig. 5.**
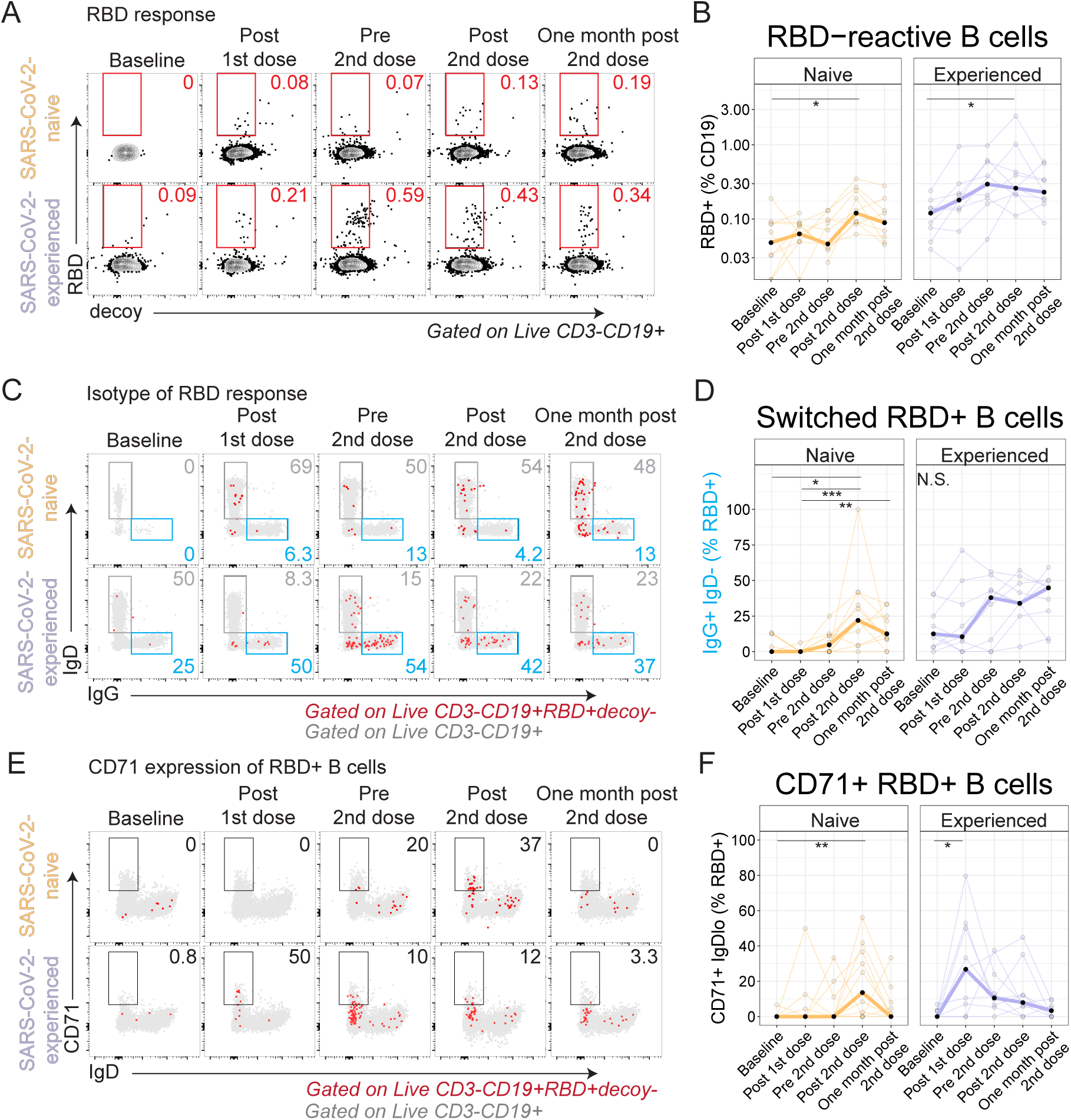
Antigen-specific B cells are induced by immunization. Recombinant biotinylated RBD was tetramerized and used to detect RBD-reactive B cells by flow cytometry. For all plots, \**P* < 0.05, \*\**P* < 0.01, and \*\*\**P* < 0.001 by Dunn’s post-test. (**A and B**) Example flow cytometry plots for RBD^+^ decoy^-^B cells (red) overlaid on all CD19^+^ B cells (grey) (A) and summary plots (B) shown. (**C and D**) RBD^+^ B cell expression of IgD and IgG (C) and summary plots (D) shown. (**E and F**) RBD^+^ B cell expression of IgD and CD71 for RBD^+^ B cells (red) overlaid on all CD19^+^ B cells (grey) (C) and summary plots (F) shown.

We next considered plasmablast differentiation, which is likely to be necessary for durable humoral immunity. Indeed, plasma cells were identified in the bone marrow of adults following COVID-19 mRNA vaccination *(52)*. Though we identified ASC (**Fig. 4**), we did not identify RBD^+^ B cells that had a CD27^+^CD38^hi^ phenotype, presumably due to down-regulation of the B cell receptor. However, other B cell subsets relevant to plasmablast differentiation were identified. For example, CD71^+^ B cells, which were described following influenza vaccination *(50)*, were induced most strongly by the second immunization in SARS-CoV-2-naive adults but were induced most strongly by the first immunization in SARS-CoV-2-experienced adults (**Fig. 5, E and F**, fig. S5D). Progressive B cell differentiation is associated with reduced expression of CD24 *(53)*. Indeed, we found that some RBD^+^ B cells had low expression of CD24 in both cohorts after vaccination (fig. S5, E and F). Similarly, B cells with low expression of CD21, which are thought to be precursors to long-lived plasma cells *(49)*, were induced, albeit weakly, in the RBD^+^ B cell subset after vaccination (fig. S5G). DN2 B cells *(54– 56)* were also increased in the RBD^+^ B cell subset after immunization (fig. S5, H and I). Together, these data indicate RBD^+^ B cells were induced by immunization and progressively developed phenotypes associated with differentiation.

### Humoral responses differ by history of COVID-19

ASC induction differed by prior history of COVID-19 (**Fig. 4**), thus we next asked whether humoral responses were affected by prior history of COVID-19. To test this, we first assessed antibody responses to the S1 subunit of the Spike protein *(57)*. As previously demonstrated *(28)*, anti-S1 IgG antibodies were detectable in individuals who had recovered from COVID-19 and were not detectable in those who were SARS-CoV-2-naive at baseline (median titers 5991 and 25, respectively; *P*=3.1×10^−8^; Wilcoxon test) (**Fig. 6A** and fig. S6A). Following first dose immunization, SARS-CoV-2-experienced adults had a median fold-change of 92 whereas SARS-CoV-2-naive adults had a median fold-change of only 2.7 (*P*=6.9×10^−4^; Wilcoxon test). However, after second dose immunization, SARS-CoV-2-experienced adults had a median fold-change of only 1.3 whereas the SARS-CoV-2-naive adults had a fold-change of 11 (*P*=3.3×10^−5^; Wilcoxon test). Compared to SARS-CoV-2-naive cohort, the SARS-CoV-2-experienced cohort had slightly higher anti-S1 IgG titers at the Post 2nd dose time point relative (*P*=0.058; Wilcoxon test; fig. S6B) but had 1.75-fold higher titers at One month post 2nd dose (*P*=5.3×10^−3^; Wilcoxon test). An overall similar pattern was observed for anti-S1 IgA titers (**Fig. 6B**), and, as expected, vaccination did not affect levels of anti-nucleocapsid antibodies (fig. S6, C to D). The change in titer in SARS-CoV-2-experienced adults was inversely correlated with their titer at baseline (fig. S6E). Thus, these data demonstrate rapid and robust humoral responses after initial vaccination in both cohorts but minimal further increase in SARS-CoV-2-experienced adults after the second vaccine dose.

**Fig. 6.**
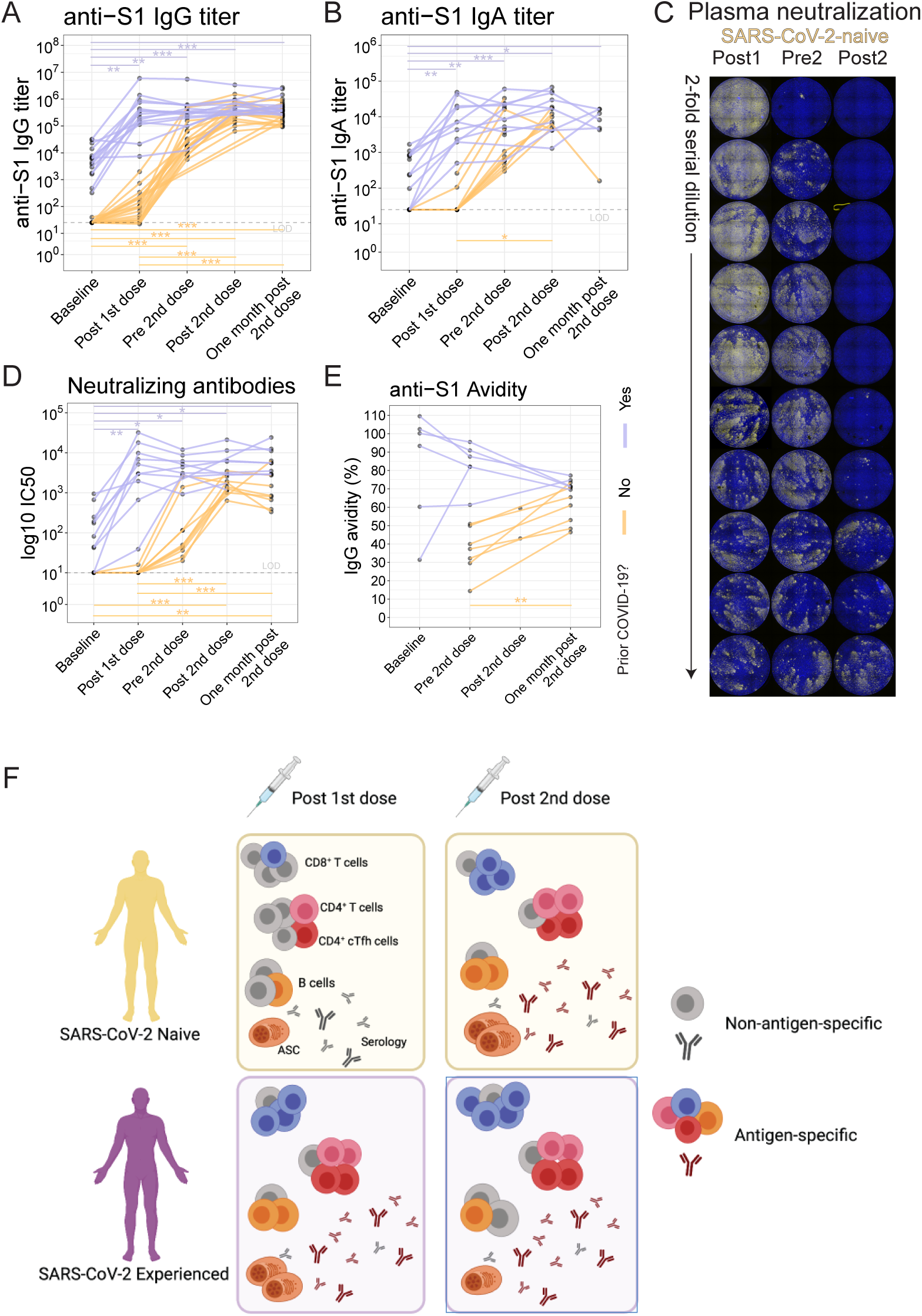
Antibody responses differ based on prior history of COVID-19. For all plots, \**P* < 0.05, \*\**P* < 0.01, and \*\*\**P* < 0.001 by Dunn’s post-test unless otherwise noted. (**A**) Anti-S1 IgG antibody titers were assessed for SARS-CoV-2 experienced (purple) and SARS-CoV-2-naive (yellow) adults. Connected lines indicate repeated measurements of the same participants over time. (**B**) Anti-S1 IgA antibody titers. (**C**) Plasma neutralizing antibody titers were assessed with an *in vitro* microneutralization assay using SARS-CoV-2 virus. Representative serial dilution series for one participant at the Post 1st dose (Post1), Pre 2nd dose (Pre2), and Post 2nd dose (Post2) time points. (**D**) Neutralizing antibody titers shown as log10 IC50. (**E**) Anti-S1 IgG antibody avidity assessed using urea wash ELISA. Data expressed as a ratio of urea washed-absorbance to unwashed absorbance. (**F**) Model for antigen specific responses for SARS-CoV-2-naive and SARS-CoV-2-experienced individuals post first and second dose.

In a subset of participants, we asked if live-virus neutralizing antibodies were induced following immunization. We observed low titers of neutralizing antibodies at baseline in SARS-CoV-2-experienced adults, whereas plasma from SARS-CoV-2-naive adults did not have detectable neutralizing antibodies (**Fig. 6, C and D**, fig. S6F). Following the first immunization, SARS-CoV-2-experienced adults had a rapid increase in neutralizing antibody titers to a median of 4084, whereas SARS-CoV-2-naive adults achieved a titer of 10 (*P*=8.7×10^−5^; Wilcoxon test). As observed earlier with anti-S1 binding antibodies, subsequent neutralizing antibody titers were largely unchanged by second vaccination in SARS-CoV-2-experienced adults, at least over the observed period, whereas the neutralizing titers in SARS-CoV-2-naive adults continued to increase. Nonetheless, at the Post 2nd dose time point, neutralizing titers remained higher in the SARS-CoV-2-experienced adults compared to the SARS-CoV-2-naive adults (fig. S6G), and also at one month post 2nd dose (*P*=2.9×10^−3^, Wilcoxon test) (fig. S6H). We also considered whether participants with a recent COVID-19 diagnosis had any effect on vaccine responses compared to participants with more distant COVID-19 diagnosis but did not find clear effects (fig. S6 I to K). In sum, we found that neutralizing antibody responses induced by SARS-CoV-2 mRNA vaccination in SARS-CoV-2-experienced adults were of greater magnitude than in naive adults.

Antibody avidity has been used to assess affinity maturation following vaccination *(58–60)*. To assess avidity, urea wash ELISA was performed for anti-S1 IgG antibodies on serum samples longitudinally. In SARS-CoV-2-naive adults, avidity continued to increase steadily over the measured time points (**Fig. 6E**), including at one month post 2nd dose when antibody titers had plateaued. This prolonged period of increasing avidity is consistent with the recent report of human axillary lymph node germinal center reactions lasting at least 5 weeks post-RNA vaccination *(21)*. All SARS-CoV-2-experienced adults assayed had relatively high-avidity antibodies at baseline, but, in contrast to SARS-CoV-2-naive adults, avidity decreased in four of five participants over time and with second vaccination, which may have been due to the induction of new, low avidity humoral responses that had not undergone germinal center maturation.

All together, these data demonstrated pronounced differences in humoral responses based on prior history of COVID-19 (**Fig. 6F**).

## DISCUSSION

Prior studies have demonstrated the importance of humoral and cellular responses for susceptibility to COVID-19 *(8)*. Better understanding of factors that affect immune responses will be critical to the design of next generation SARS-CoV-2 vaccines and their optimal use, including booster shots. Here, we observed subtle differences in cellular responses and more pronounced differences in humoral responses between individuals naive to SARS-CoV-2 and those who had recovered from SARS-CoV-2 infection. Both cohorts had similar robust activated CD8^+^ T cell responses to vaccination, which were typified by co-expression of Ki67 and CD38, whereas activated CD4^+^ T cell responses were generally more muted. Among CD4^+^ responses, we found evidence of antigen-specific CD4^+^ and CD8^+^ T cell responses. Furthermore, antigen-specific B cell responses differed by cohort as well. SARS-CoV-2-experienced adults had more antigen-specific ASC in circulation one week after the first vaccination compared to SARS-CoV-2-naive adults, but the frequency of antigen-specific ASC after second vaccination did not increase in previously infected individuals, unlike the SARS-CoV-2-naive adults. In both cohorts, however, RBD-specific B cells increased in frequency in circulation following vaccination. Additionally, prior history of COVID-19 was associated with 100-to 1000-fold increase in anti-Spike IgG antibody titers following the first vaccination, with limited increase upon the second vaccination, whereas antibody titers increased steadily over time in SARS-CoV-2-naive adults.

Prior studies demonstrated potent induction of T cell responses in animal models *(33)*. Consistent with these studies, here we also observed robust induction of cytotoxic CD8^+^ T cell responses following vaccination. The Ki67^+^CD38^+^ CD4^+^ T cell responses were muted but were consistent with other reports that have identified antigen-specific CD4^+^ T cell responses following mRNA vaccination *(14)*, thus indicating establishment of CD4^+^ T cell responses following vaccination. Moreover, we demonstrated the induction of Spike-specific responses in both CD8^+^ and CD4^+^ T cells among SARS-CoV-2-naive adults using AIM. Although a robust CD8^+^Ki67^+^CD38^+^ T cell response was observed phenotypically in PBMC from SARS-CoV-2-experienced adults, the AIM responses were low magnitude. Further optimization of the peptide pools may be needed, as other studies have observed strong peptide-specific CD8^+^ responses. Additional studies of antigen specificity, such as by T cell tetramer assays, may help resolve T cell responses to individual epitopes. Future studies will be needed to determine whether Spike-specific T cell responses after mRNA immunization in adults are a correlate of protection.

Durable, affinity-matured antibody responses require germinal center reactions in lymphoid tissue *(42)*, and germinal centers are indeed established following mRNA vaccination *(21)*. Consistent with this, our Spike antibody avidity experiments demonstrate progressive maturation of serum antibodies following mRNA vaccination, with increasing avidity through at least a month post second vaccination. However, much remains to be learned about Tfh responses in this context and whether these cells play a role in poor antibody responses after mRNA vaccination *(61)*. Two prior studies evaluated mRNA vaccination for influenza in humans and non-human primates and found robust induction of ICOS^+^CXCR3^+^PD-1^+^ cTfh responses and neutralizing antibodies *(48, 62)*. Here, CXCR3^+^ICOS^+^CD38^+^ cTfh were induced by vaccination in both cohorts. Furthermore, we previously demonstrated that antigen-specific cTfh responses were induced by influenza vaccination which correlated with cellular and humoral responses to vaccination *(38, 63)*. Here as well, we found that mRNA vaccination induced Spike-specific activated cTfh. However, activated cTfh are, at best, a proxy for understanding lymphoid Tfh responses in humans *(43, 63)*. Plasma CXCL13, which increased after yellow fever vaccination but not influenza vaccination *(51, 64)*, was not different following mRNA vaccination in our study, which may indicate relative differences in extent of germinal center formation or perhaps differences in early germinal center events that lead to CXCL13 production. Future studies involving direct lymph node sampling, as was done recently *(65, 66)*, will be needed to understand Tfh dynamics and memory formation, and additional time points will be needed to determine the utility of cellular and humoral biomarkers, including plasma CXCL13 and the plasmablast response, following mRNA vaccination.

Notable qualitative and quantitative differences in immune responses were observed when comparing adults who were naive to SARS-CoV-2 to those who had recovered from SARS-CoV-2 infection. Anti-S1 binding IgG antibodies and neutralizing antibodies appeared to peak one week after second vaccination, consistent with published reports of humoral responses to mRNA vaccination *(12, 18)*. By the end of the observation period, SARS-CoV-2-experienced adults had higher titers of anti-S1 binding IgG and neutralizing antibodies at the One month post 2nd dose time point, compared to SARS-CoV-2-naive adults. Moreover, humoral responses continued to qualitatively change in avidity for Spike protein despite the plateau in antibody quantity. In SARS-CoV-2-naive adults, affinity increased over time, which may reflect germinal center-related affinity maturation *(11, 67)*. In contrast, SARS-CoV-2-experienced adults had reduction in affinity over time, presumably reflecting the contribution of *de novo* B cell responses that had not undergone affinity maturation, rather than loss of high-affinity antibodies. Indeed, better understanding the qualitative and quantitative changes in antibodies over time will have major implications for the need for booster vaccinations.

Furthermore, humoral responses were robust after the first vaccination but more muted after the second vaccination in SARS-CoV-2-experienced adults, and this pattern was also evident in anti-S1 IgG and anti-S1 IgA antibodies, as well as live-virus neutralizing antibodies. Several possibilities may explain these differences. For example, the early plateau in humoral responses could indicate altered B cell differentiation away from antigen-specific plasma cells, which would be consistent with the relatively poor ASC responses in SARS-CoV-2-experienced adults after the second dose. In addition, the reduction in antigen-specific ASC may have also altered trafficking of ASC following repeat vaccination, perhaps shifting the peak ASC response earlier than was assessed here. Another possibility is that the very high titers of anti-S1 IgG responses may restrict antigen availability for stimulation of non-memory B cell clones following subsequent vaccine doses. Indeed, there was a strong negative correlation between the baseline anti-S1 IgG titer and the fold-change in anti-S1 IgG titers after first vaccination. Furthermore, differences between cohorts could arise from other differences prior to lymphocyte activation, such as during antigen-presenting cell priming, as the duration of the dysregulation of innate immune responses in the setting of COVID-19 remains unknown *(68)*. Future studies will be needed to better understand adaptive immune responses to COVID-19 mRNA vaccination, which will have direct implications for durable, effective protection from infection, and the need for boosters.

Together, these results highlight the importance of understanding prior immunological experience on the subsequent immune response to COVID-19 mRNA vaccines. Future studies will be needed to determine whether such personalized vaccination regimens will deliver durable, protective immunity to infection by the SARS-CoV-2 virus.

## MATERIALS AND METHODS

### Study Design

Thirty-six adults (21 SARS-CoV-2-naïve and 15 SARS-CoV-2-experienced) provided written consent for enrollment with approval from the NYU Institutional Review Board (protocols 18-02035 and 18-02037).

### Blood samples processing and storage

Venous blood was collected by standard phlebotomy. Blood collection occurred at baseline, approximately one week after first vaccination (“Post 1st dose”), prior to the second vaccination (“Pre 2nd dose”), approximately one week after the second vaccination (“Post 2nd dose”), and one month after the second vaccination (“One month post 2nd dose”), as depicted in **Fig. 1A**. Peripheral blood mononuclear cells (PBMC) were isolated from heparin vacutainers (BD Biosciences) that were stored overnight at room temperature (RT), followed by processing using Sepmates (Stem Cell, Inc) in accordance with the manufacturer’s recommendations. Serum was collected in SST tubes (BD Biosciences) and frozen immediately at -80°C.

### ELISA

Direct ELISA was used to quantify antibody titers in participant serum. Ninety-six well plates were coated with 1 µg/mL S1 protein (100 µL/well) or 0.1 µg/mL N protein diluted in PBS and were then incubated overnight at 4°C (Sino Biological Inc., 40591-V08H and 40588-V08B). Plates were washed four times with 250 µL of PBS containing 0.05% Tween 20 (Fisher) (PBS-T) and blocked with 200 µl PBS-T containing 4% non-fat milk and 5% whey, as blocking buffer at RT for 1 hour. Sera were heated at 56°C for 1 hour prior to use. Samples were diluted to a starting concentration of 1:50 (S1), or 1:100 (N) were first added to the plates and then serially diluted 1:3 in blocking solution. The final volume in all wells after dilution was 100 µL. After a 2-hour incubation period at RT, plates were washed four times with PBS-T. Horseradish-peroxidase conjugated goat-anti human IgG, IgM, and IgA (Southern BioTech, 2040-05, 2020-05, 2050-05) were diluted in blocking buffer (1:2000, 1:1000, 1:1000, respectively) and 100 µL was added to each well. Plates were incubated for 1 hour at RT and washed four times with PBS-T. After developing for 5 min with TMB Peroxidase Substrate 3,3′,5,5′-Tetramethylbenzidine (Thermo Scientific), the reaction was stopped with 1M sulfuric acid or 1N hydrochloric acid. The optical density was determined by measuring the absorbance at 450 nm on a Synergy 4 (BioTek) plate reader.

In order to summarize data collected on individuals, the area under the response curve was calculated for each sample and end point titers were normalized using replicates of pooled positive control sera on each plate to reduce variability between plates.

### Avidity assay

Ninety-six well plates were coated with 0.1 µg/mL S1 protein (100 µL/well) diluted in PBS overnight at 4°C (Sino Biological). Plates were washed four times with 250 µL of PBS containing 0.05% Tween 20 (PBS-T) and blocked with 200 µL PBS-T containing 4% non-fat milk and 5% whey, as blocking buffer at RT for 1 hour. Sera were heated at 56°C for 1 hour prior to use. Samples were diluted to a starting concentration of 1:50 and added to the plates in quadruplicate and then serially diluted 1:3 in blocking solution. The final volume in all wells after dilution was 100 µL. After a 2-hour incubation period at RT, plates were washed four times with PBS-T. PBS was then added to two dilution replicate sets and 6 M Urea to the other two dilution replicate sets. Plates were incubated for 10 min at RT before washing four times with PBS-T. Antibodies were detected and plates were developed and read as described above for ELISA assays.

Avidity was calculated by dividing the dilutions that gave an optical density value of 0.5 (Urea treatment/no Urea). Scores with theoretical values between 0 and 100% were generated.

### Antibody-secreting cell ELISpot Assays

A direct enzyme-linked immunospot (ELISpot) assay was used to determine the number of SARS-CoV-2 spike protein subunit S1-, S2-, and receptor-binding domain (RBD)-specific IgG, IgA, and IgM ASCs in fresh PBMCs. Ninety-six well ELISpot filter plates (Millipore, MSHAN4B50) were coated overnight with 2 µg/mL recombinant S1, S2, or RBD (Sino Biological Inc., 40591-V08H, 40590-V08B, and 40592-V08H), or 10 µg/mL of goat anti-human IgG/A/M capture antibody (Jackson ImmunoResearch Laboratory Inc., 109-005-064). Plates were washed 4 times with 200 µL PBS-T and blocked for 2 hours at 37°C with 200 µL RPMI 1640 containing 10% fetal calf serum (FCS), 100 units/mL of penicillin G, and 100 μg/mL of streptomycin (Gibco), referred to as complete medium. Fifty µL of cells in complete media at 10×10^6^ cells/mL were added to the top row of wells containing 150 µL complete media and 3-fold serial diluted 3 times. Plates were incubated overnight at 37°C with 5% CO_2_. Plates were washed 1 time with 200 µL PBS followed by 4 times with 200 µL PBS containing 0.05% Tween 20 (PBS-T). Biotinylated anti-human IgG, IgM, or IgA antibody (Jackson ImmunoResearch Laboratory Inc., 709-065-098, 109-065-129, 109-065-011) were diluted 1:1000 in PBS-T with 2% FCS (Ab diluent) and 100 µL was added to wells for 2 hours at RT or overnight at 4°C. Plates were washed four times with 200 µL PBS-T and incubated with 100 µL of Avidin-D-HRP conjugate (Vector Laboratories, A-2004) diluted 1:1000 in PBS-T for 1 hour at RT. Plates were washed 4 times with 200 µL PBS-T and 100 mL of AEC substrate (3 amino-9 ethyl-carbazole; Sigma Aldrich, A-5754) was added. Plates were incubated at RT for five minutes and rinsed with water to stop the reaction. Developed plates were scanned and analyzed using an ImmunoSpot automated ELISpot counter (Cellular Technology Limited).

### SARS-CoV-2 microneutralization assay

Viral neutralization activity of plasma was measured in an immunofluorescence-based microneutralization assay by detecting the neutralization of infectious virus in cultured Vero E6 cells (African Green Monkey Kidney; ATCC #CRL-1586). These cells are known to be highly susceptible to infection by SARS-CoV-2. Cells were maintained according to standard ATCC protocols. Briefly, Vero E6 cells were grown in Dulbecco’s Modified Eagle Medium (DMEM) supplemented with 10% heat-inactivated fetal bovine serum (FBS), 2 mM L-glutamine, and 1% of MEM Nonessential Amino Acid (NEAA) Solution (Fisher #MT25025CI). Cell cultures were grown in 75 or 150 cm^2^ flasks at 37°C with 5% CO_2_ and passaged 2-3 times per week using trypsin-EDTA. Cell cultures used for virus testing were prepared as subconfluent monolayers. All incubations containing cells were performed at 37°C with 5% CO_2_. All SARS-CoV-2 infection assays were performed in the CDC/USDA-approved BSL3 facility of NYU Grossman School of Medicine (New York, NY), in accordance with its Biosafety Manual and Standard Operating Procedures. SARS-CoV-2 isolate USA-WA1/2020, deposited by the Centers for Disease Control and Prevention, was obtained through BEI Resources, NIAID, NIH (NR-52281, GenBank accession no. MT233526). Serial dilutions of heat-inactivated plasma (56°C for 1 hour) were incubated with USA-WA1/2020 stock (at fixed 1×10^6^ PFU/mL) in DMEM supplemented with 2 mM L-glutamine, 1% of MEM Nonessential Amino Acid (NEAA) Solution, and 10 mM Hepes (ThermoFisher 15-630-080) for 1 hour at 37°C. One hundred microliters of the plasma-virus mix was then added to the cells and incubated at 37°C with 5% CO_2_. Twenty-four hours post-infection, cells were fixed with 10% formalin solution (4% active formaldehyde) for 1 hour, stained with an α-SARS-CoV-2 nucleocapsid antibody (ProSci #10-605), and a goat α-mouse IgG AF647 secondary antibody along with DAPI and visualized by microscopy with the CellInsight CX7 High-Content Screening (HCS) Platform (ThermoFisher) and high-content software (HCS) *(69)*.

### CXCL13 detection

CXCL13 was detected using the Ella instrument (ProteinSimple) and a CXCL13 Simple Plex Cartridge (ProteinSimple, SPCKB-PS-000375) on serum diluted 1:1 in buffer according to the manufacturer’s instruction.

### Cellular phenotyping

Peripheral blood was collected in sodium heparin collection tubes and maintained at room temperature overnight. PBMC were isolated using the Sepmate system (STEMCELL Technologies) in accordance with manufacturer’s instructions. Then, 2 to 5 million freshly isolated PBMC were resuspended in HBSS supplemented with 1% fetal calf serum (Fisher) and 0.02% sodium azide (Sigma). Cells underwent Fc-blockade with Human TruStain FcX (Biolegend) and NovaBlock (ThermoFisher) for 10 minutes at room temperature, followed by surface staining antibody cocktail at room temperature for 20 minutes in the dark. Cells were permeabilized with the eBioscience Intracellular Fixation and Permeabilization kit (Fisher) for 20 minutes at room temperature in the dark, followed by intracellular staining with an antibody cocktail for 1 hour at room temperature in the dark. All samples were then resuspended in 1% paraformaldehyde and acquired within three days of staining on a 5-laser Aurora cytometer (Cytek Biosciences). Antibodies, clones, and catalog numbers are described in **Table S3**. Initial data quality control was performed using FlowJo. Non-naive CD8^+^ and CD4^+^ T cells were analyzed in the OMIQ.ai platform (www.omiq.ai) using Phenograph clustering *(32)* with k=20 and a Euclidean distance metric, followed by tSNE projection. Heatmaps and differential cluster abundance were assessed by edgeR *(70)* via OMIQ.ai.

### Activation-induced marker analysis

Cryopreserved PBMC were thawed and rested overnight at 37C in RPMI 1640 with L-glutamine (Fisher) containing 10% FCS (Fisher), 2 mM L-glutamine (Fisher), and 100 U/ml of penicillin-streptomycin (Fisher). The following day, cells were stimulated with 0.6 nmol of each of the S1, S, and S+ PepTivator pools (Miltenyi) for 20 hours at 37C with 1.5×10^6^ cells per well in a 96 well flat bottom plate. For the unstimulated control well, sterile water was used in place of the peptide pools. Monensin (ThermoFisher) was added for the last 6 hours of stimulation at a final concentration of 10 uM. After stimulation, cells were washed with PBS containing 10 mM EDTA at 37°C for 5 minutes, followed by Fc-blockade and were stained as previously described. Antibodies, clones, and catalog numbers are described in **Table S3**. Analysis was performed using FlowJo.

### B cell tetramer assays

Recombinant biotinylated RBD (Biolegend) was reacted with PE-Streptavidin (Biolegend) in a 4:1 molar ratio at 4C for 2 hours. For flow cytometry studies, PBMC underwent Fc-blockade with Human TruStain FcX (Biolegend), NovaBlock (Fisher), APC-Streptavidin (Biolegend), and mouse PE-IgG2b isotype control (Biolegend) for 10 minutes at room temperature, followed by APC anti-PE (Biolegend) for 10 minutes. Then cells were washed twice and stained with the RBD tetramer and antibodies against other surface proteins for 20 minutes at room temperature in the dark, followed by resuspension in 1% paraformaldehyde and acquisition on a 5-laser Aurora cytometer (Cytek Biosciences).

### Bioinformatics and statistical analyses

Primary data analysis and statistical analysis were performed using the R environment (version 4.0.2) and all bioinformatics scripts are available at https://github.com/teamTfh/COVIDvaccines. Statistical tests were performed using the “rstatix” library (version 0.6.0). Nonparametric 2-sample *t*-tests were performed using Wilcoxon tests where sample variances were unequal, as identified by Levene’s test. Use of parametric or nonparametric tests was guided by Shapiro-Wilk normality testing and Bartlett’s test for homoscedasticity. Parametric ANOVA were performed with Tukey’s post-test, whereas non-parametric ANOVA were performed as Kruskal-Wallis tests with Dunn’s post-test. Correlation analyses were performed as nonparametric tests using Kendall’s tau statistic. All tests were two-tailed tests with α=0.05. Study schematics were made with BioRender.

## Data Availability

Data from this study is available upon request.

## Supplementary Materials

Fig. S1. CD4 and CD8 T cell responses and gating strategy.

Fig. S2. Antigen-specific T cell responses to vaccination.

Fig. S3. Plasmablasts and CXCL13 responses to vaccinations.

Fig. S4. Poor IgG and IgA ASC responses to second dose in SARS-CoV-2 experienced participants.

Fig. S5. RBD-reactive B cells following vaccination.

Fig. S6. SARS-CoV-2-experienced individuals’ robust anti-S1 binding and neutralizing antibodies responses after the first dose.

Table S1. Participant demographics.

Table S2. Clinical COVID-19 disease among SARS-CoV-2-experienced participants

Table S3. Antibodies used in flow cytometry experiments.

## Acknowledgements

We would like to thank the participants who joined our studies. We would like to thank all of the NYU Vaccine Center staff, including Juanita Erb, Gali Moritz, Mahnoor Ali, Stephanie Rettig, Heekoung Youn, Brooklyn Henderson, Lisa Zhao, and Harry Lambert for their help in these studies. We thank Maren De Vries for the viral stock production and titration. We thank Meike Dittmann and The Microscopy Laboratory at NYU Langone Health for the use of their microscopes. We thank the Office of Science & Research High-Containment Laboratories at NYU Grossman School of Medicine for their support in the completion of this research and Ludovic Desvignes for his management of the NYU Langone Biosafety Level 3 laboratory. We also thank the research clinicians at the NYU Langone Vaccine Center Manhattan clinic for assistance with study participant visits including: Tamia Davis, Vanessa Raabe, Purvi Parikh, Alexander McMeeking, Rebecca Pellet-Maddan, and Bo Shopsin.

## Author contributions

Conceptualization: M.I.S., A.C., R.S.H., and M.J.M.

Data curation: M.I.S., A.C., S.L.G-G., and R.S.H.

Formal analysis: M.I.S., A.C., S.L.G-G., S.B.K., and R.S.H.

Funding acquisition: M.J.M., and R.S.H.

Investigation: M.I.S, A.C., J.R.A, S.L.G-G., T.K., J.P.W., S.W.H., M.T., R.S.H.

Methodology: M.I.S., A.C., and R.S.H.

Project administration: M.I.S. and R.S.H.

Supervision: R.S.H. and M.J.M.

Validation: M.I.S., S.L.G-G, A.C., and R.S.H.

Visualization: R.S.H.

Writing – original draft: M.I.S. and R.S.H.

Writing – review & editing: M.I.S., A.C., S.L.G-G., S.B.K.,J.P.W., T.K., M.T., M.J.M., S.H., and R.S.H.

