## Supplemental Figures and Tables for "Robust immune responses after one dose of BNT162b2 mRNA vaccine dose in SARS-CoV-2 experienced individuals"

### Supplementary Materials

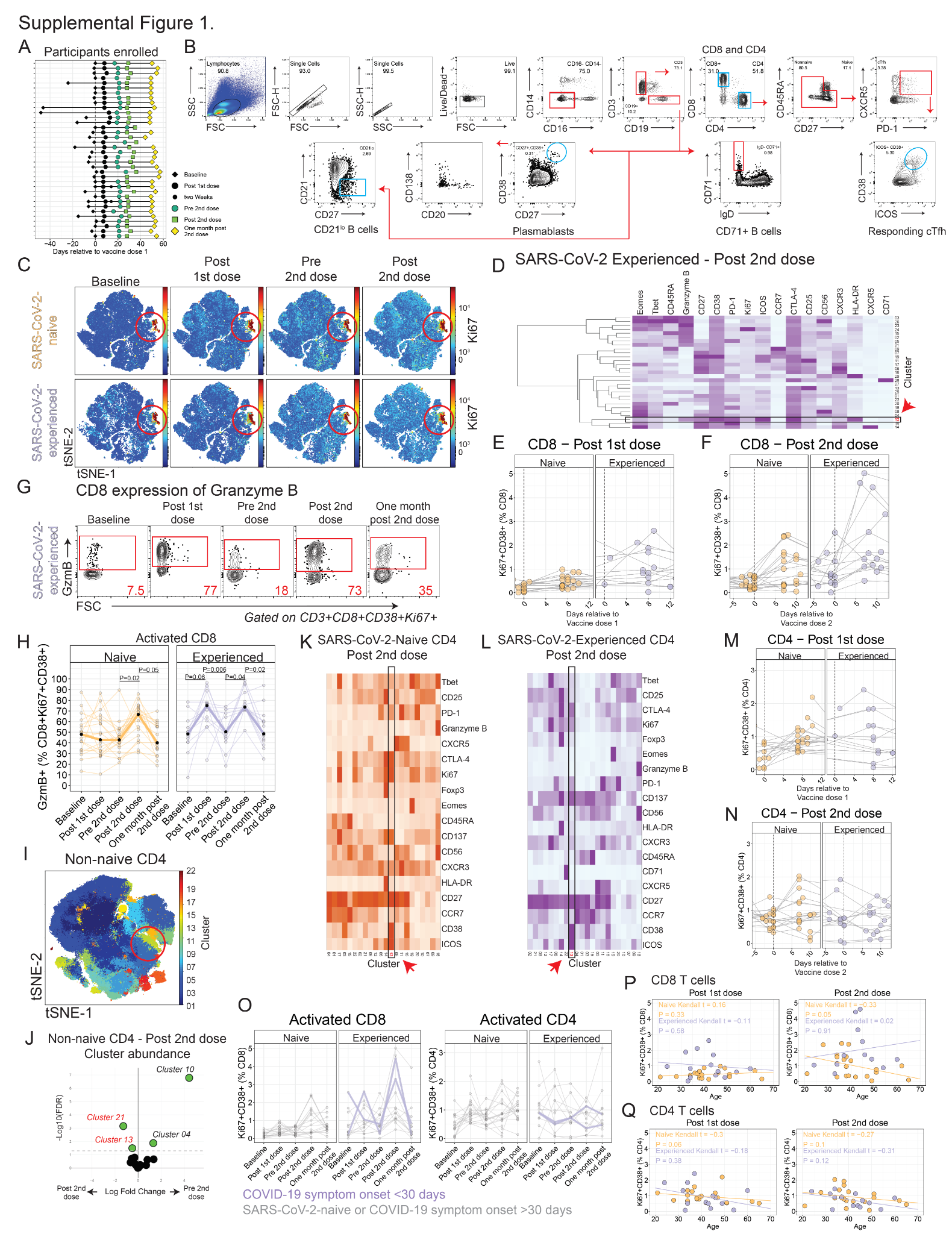

#### Fig. S1. CD4^+^ and CD8^+^ T cell responses and gating strategy.

(**A**) Study participant timeline relative to first vaccination. (**B**) Gating scheme for T and B cell populations. (**C**) Non-naive CD8 shown in tSNE projection for SARS-CoV-2-naive (upper) or SARS-CoV-2-experienced (lower) participants. Heatmap shows expression of Ki67. Circled area indicates region corresponding to Cluster 12. (**D**) Non-naive CD8^+^ underwent Phenograph clustering. Protein expression for each cluster for SARS-CoV-2-experienced adults shown for samples taken one week after the second vaccination. (**E** and **F**) Ki67^+^CD38^+^ expression in CD8 T cells by cohort over time measured in days, relative to the individual’s first (E) or second (F) vaccination. (**G**) Example for Ki67^+^CD38^+^ CD8^+^ T cell expression of granzyme B in a SARS-CoV-2-experienced individual. Red number indicates frequency. (**H**) Summary data for Ki67^+^CD38^+^ CD8^+^ T cell expression of granzyme B. *P*-values from one-way ANOVA with Tukey’s post test. (**I**) Non-naive CD4^+^ T cells from all samples were merged for tSNE projection. Colors indicate Phenograph clustering. (**J**) Phenograph cluster abundance for non-naive CD4^+^ T cells for all participants before and after second vaccination. (**K and** **L**) Protein expression for Phenograph clusters for non-naive CD4^+^ T cells shown for samples at one week following second vaccination in SARS-CoV-2-naive (K) or SARS-CoV-2-experienced (L) participants. (**M and N**) CD4^+^ T cells shown for expression of Ki67 and CD38 after vaccination over time measured in days, relative to the individual’s first (M) or second (N) vaccinations. (**O**) Summary data for Ki67^+^CD38^+^ expression in CD8^+^ T cells (left) and CD4^+^ T cells (right) by cohort highlighting the two recent COVID infection participants in blue. (**P and Q**) Correlation between Ki67^+^CD38^+^ CD4^+^ (P) or CD8^+^ (Q) T cells and age one after either first vaccination (left) or second vaccination (right).

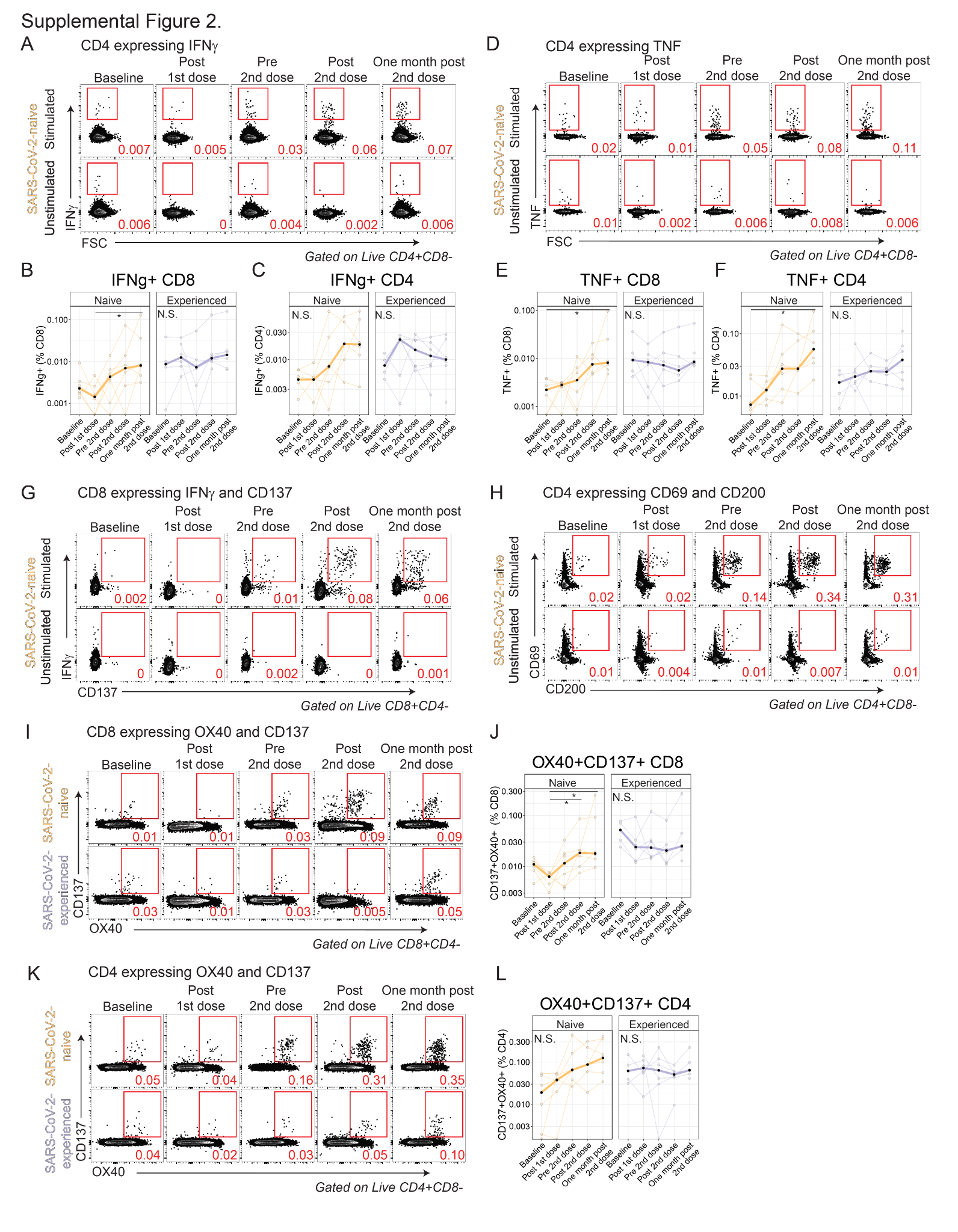

#### Fig. S2. Antigen-specific T cell responses to vaccination.

PBMC were rested overnight, then stimulated for 20 hours with SARS-CoV-2 Spike peptides in the presence of monensin, followed by phenotyping for flow cytometry. For all panels, **P* < 0.05 by parametric or non-parametric ANOVA with multiple comparisons correction. (**A**) CD4^+^ T cells shown for expression of IFNγ in a SARS-CoV-2-naive individual with peptide stimulation (top row) or no stimulation (bottom row). (**B and C**) Summary plots for expression of IFNγ in CD8^+^ (B) or CD4^+^ (C) T cells. (**D**) CD4^+^ T cells shown for expression of TNF in a SARS-CoV-2-naive individual with peptide stimulation (top row) or no stimulation (bottom row). (**E and F**) Summary plots for expression of TNF in CD8^+^ (E) or CD4^+^ (F) T cells. (**G**) CD8^+^ T cell expression of IFNγ and CD137 shown for one representative SARS-CoV-2-naive individual across all time points for stimulated (top row) or unstimulated (bottom row) conditions. (**H**) CD4^+^ T cell expression of CD69 and CD200 for stimulated and unstimulated conditions. (**I to L**) Expression of OX40 and CD137 in CD8^+^ (I) or CD4^+^ (K) T cells in SARS-CoV-2-naive (top row) and SARS-CoV-2-experienced (bottom row) individuals, with summary plots for CD8^+^ (J) and CD4^+^ (L) at right. Red number indicates frequency.

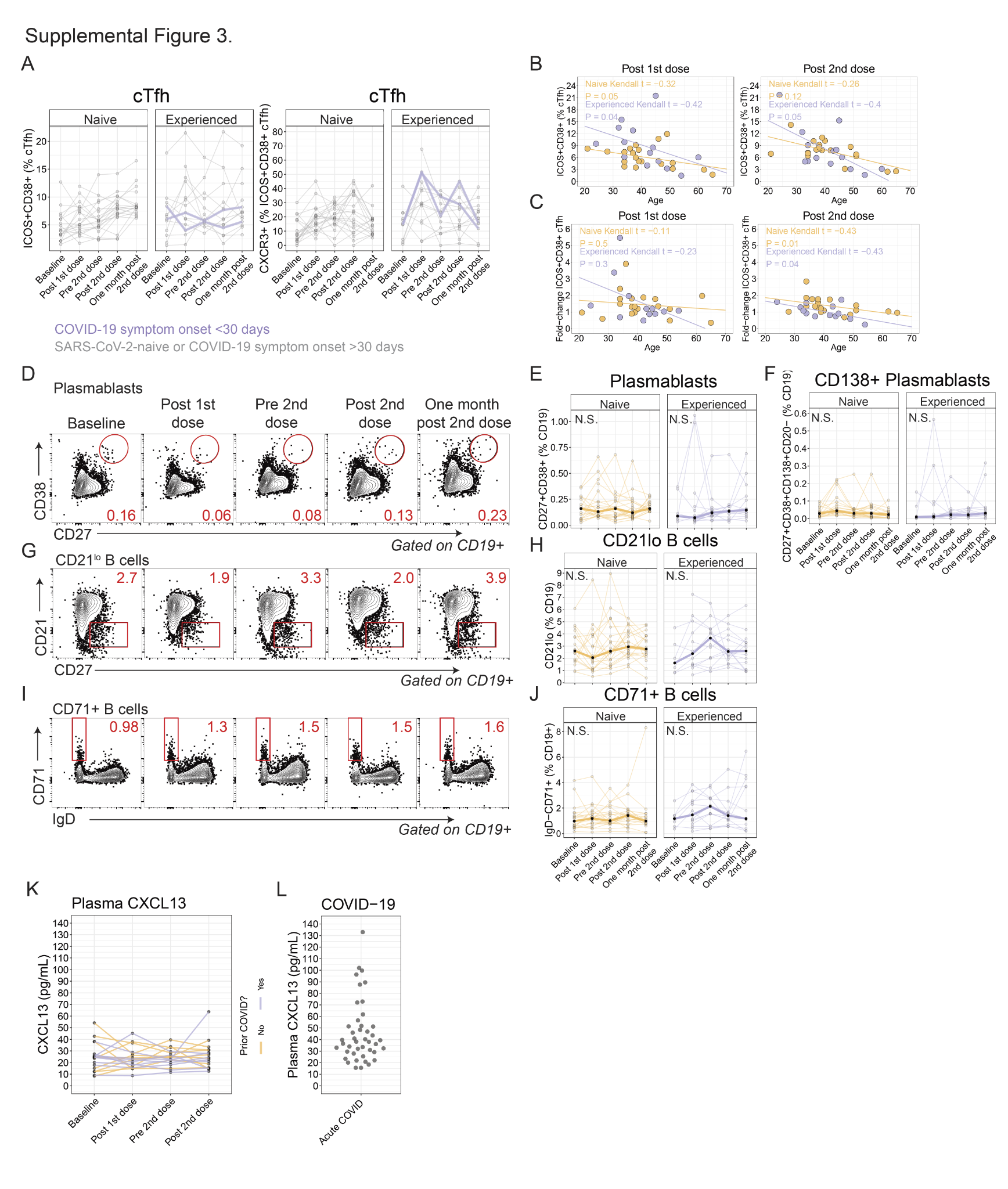

#### Fig. S3. Plasmablast and CXCL13 responses to vaccinations.

(**A**) Summary data for expression of ICOS and CD38 in cTfh (left) and CXCR3 expression in ICOS^+^CD38^+^ cTfh (right) by cohort highlighting the two recent COVID infection participants in blue. (**B**) cTfh expressing ICOS and CD38 were negatively correlated with age one week after the first vaccination (left) or the second vaccination (right). (**C**) The fold-change in cTfh expressing ICOS and CD38 at one week after the first vaccination compared to baseline (left) or 1 week after the second vaccination compared to Pre 2nd dose (right) was negatively correlated with age. (**D**) Plasmablasts were identified by expression of CD27 and CD38. Example plot shown. (**E**) Summary data for plasmablasts identified by high expression of CD27 and CD38. (**F**) Summary data shown for plasma cells, defined as CD27^+^CD38^hi^CD138^+^CD20^-^, as a proportion of CD19^+^ B cells. (**G**) Example plots showing gating for CD21^lo^ B cells. (**H**) Summary data for CD21^lo^ B cells longitudinally. (**I**) Example plots showing gating for CD71^+^ B cells. Red number indicates frequency. (**J**) Summary data for CD71^+^ B cells longitudinally. (**K**) Plasma CXCL13 shown longitudinally for both cohorts. (**L**) Plasma CXCL13 in an independent cohort of patients with acute COVID-19 who were sampled 30 days of the onset of symptoms.

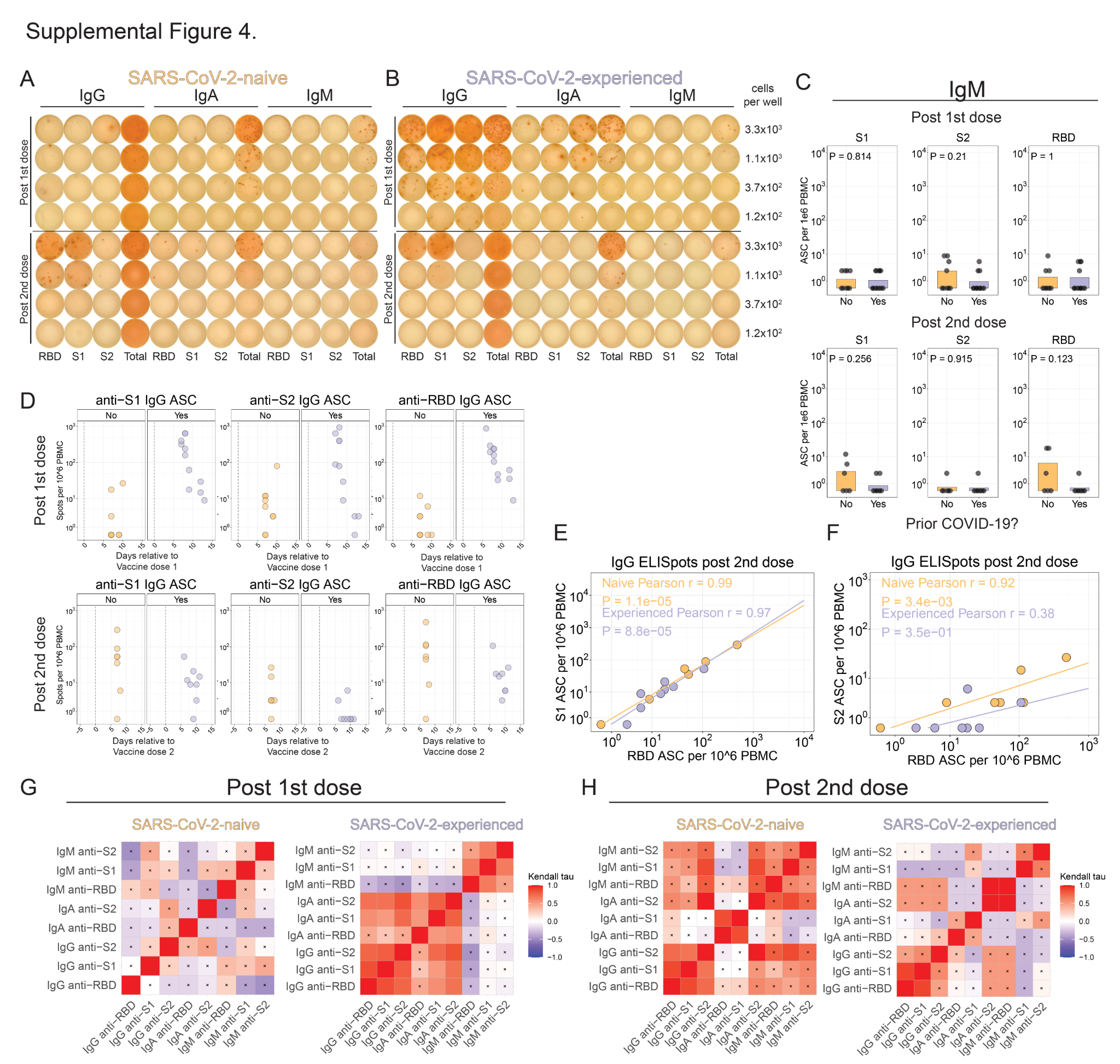

#### Fig. S4. Poor IgG and IgA ASC responses to second dose in SARS-CoV-2 experienced participants.

(**A and** **B**) Antibody-secreting cell (ASC) ELISpots shown for IgG, IgA, and IgM-producing cells reacting to RBD, S1, or S2 antigens, or total secreted antibody controls. (**C**) IgM-producing ASC in circulation quantified one week after first vaccination (top) or one week after second vaccination (bottom). Nominal *P* values from Wilcoxon tests. (**D**) ASC frequencies shown over time measured in days relative to the first vaccination (top) or second vaccination (bottom) for S1, S2, or RBD antigens. (**E** **and F**) Correlation shown for both cohorts for S1-reactive IgG ASC (E) or S2-reactive IgG ASC (F) compared to RBD-reactive IgG ASC one week after second vaccination. (**G and** **H**) Kendall correlation shown for SARS-CoV-2-specific frequencies for SARS-CoV-2-naive or SARS-CoV-2-experienced adults one week after first (G) or one week after second (H) vaccination. Heatmap colored by Kendall’s tau statistic. Boxes with symbols indicate nominal *P* value >0.05.

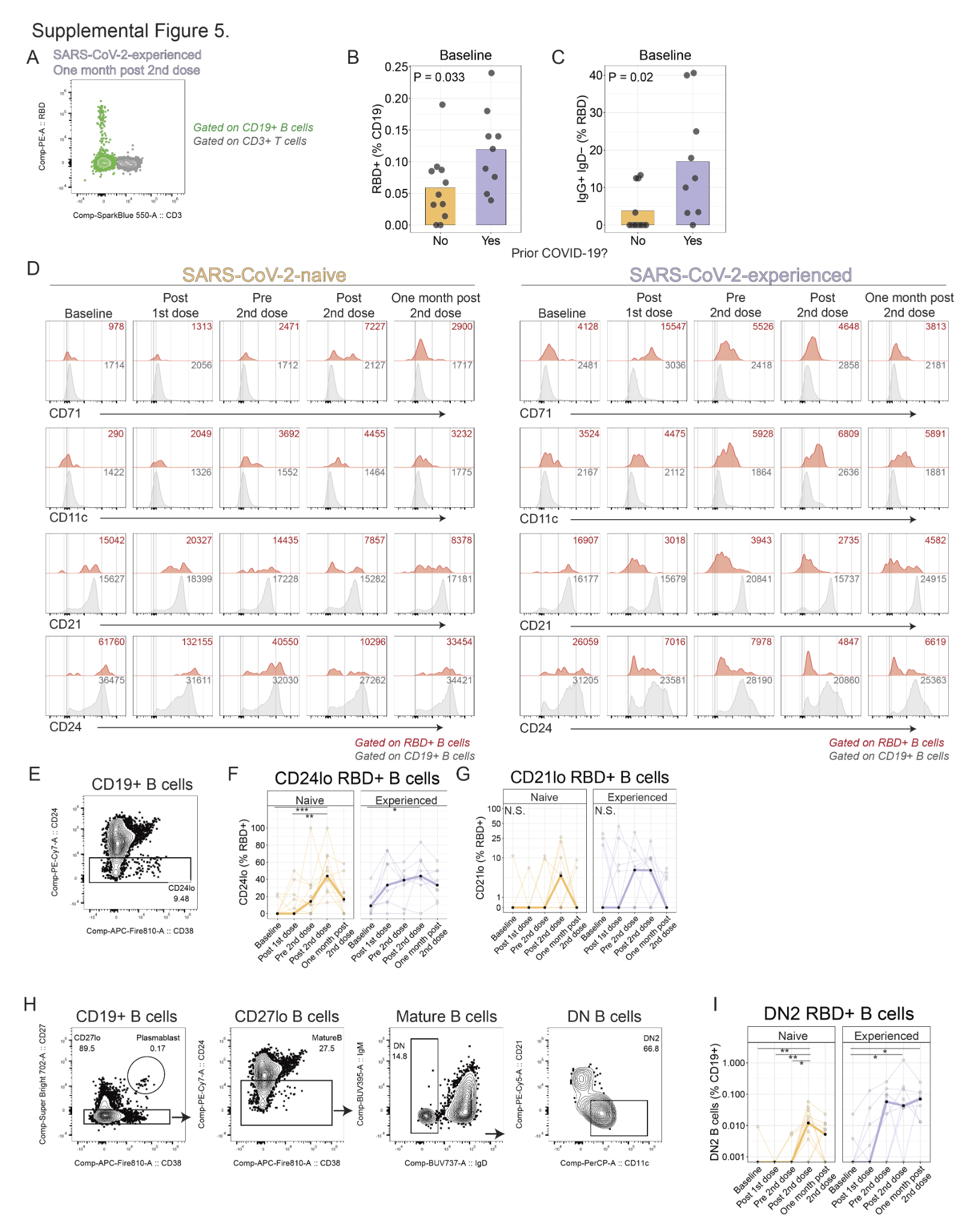

#### Fig. S5. RBD-reactive B cells following immunization.

(**A**) Recombinant biotinylated RBD was tetramerized using PE-streptavidin. A SARS-CoV-2-experienced individual late after vaccination was used as an initial positive control. Plot shows CD19^+^ B cells (green) or CD3^+^ T cells (gray) for RBD binding. (**B**) RBD-reactive B cells were compared at baseline in SARS-CoV-2-naive (orange) or SARS-CoV-2-experienced (purple) participants (*P*=0.03, Wilcoxon test). (**C**) Proportion of IgG-expressing B cells among all RBD^+^ B cells by cohort (*P*=0.02, Wilcoxon test). (**D**) B cell phenotypic analysis shown. For each row, the red histogram depicts RBD^+^ B cells and the grey histogram depicts all CD19^+^ B cells, with the numbers indicating the mean fluorescence intensity for the respective populations for the proteins shown in each row. (**E**) Gating scheme for identification of CD24^lo^ B cells. (**F and G**) Summary plots for CD21^lo^ B cells (F) and CD24^lo^ B cells (G) among all RBD^+^ B cells. (**H**) Gating scheme for identification of DN2 B cells. (**I**) Summary plots for proportions of DN2 B cells. * *P* < 0.05, ** *P* < 0.01, and ****P* < 0.001 by Dunn’s post-test.

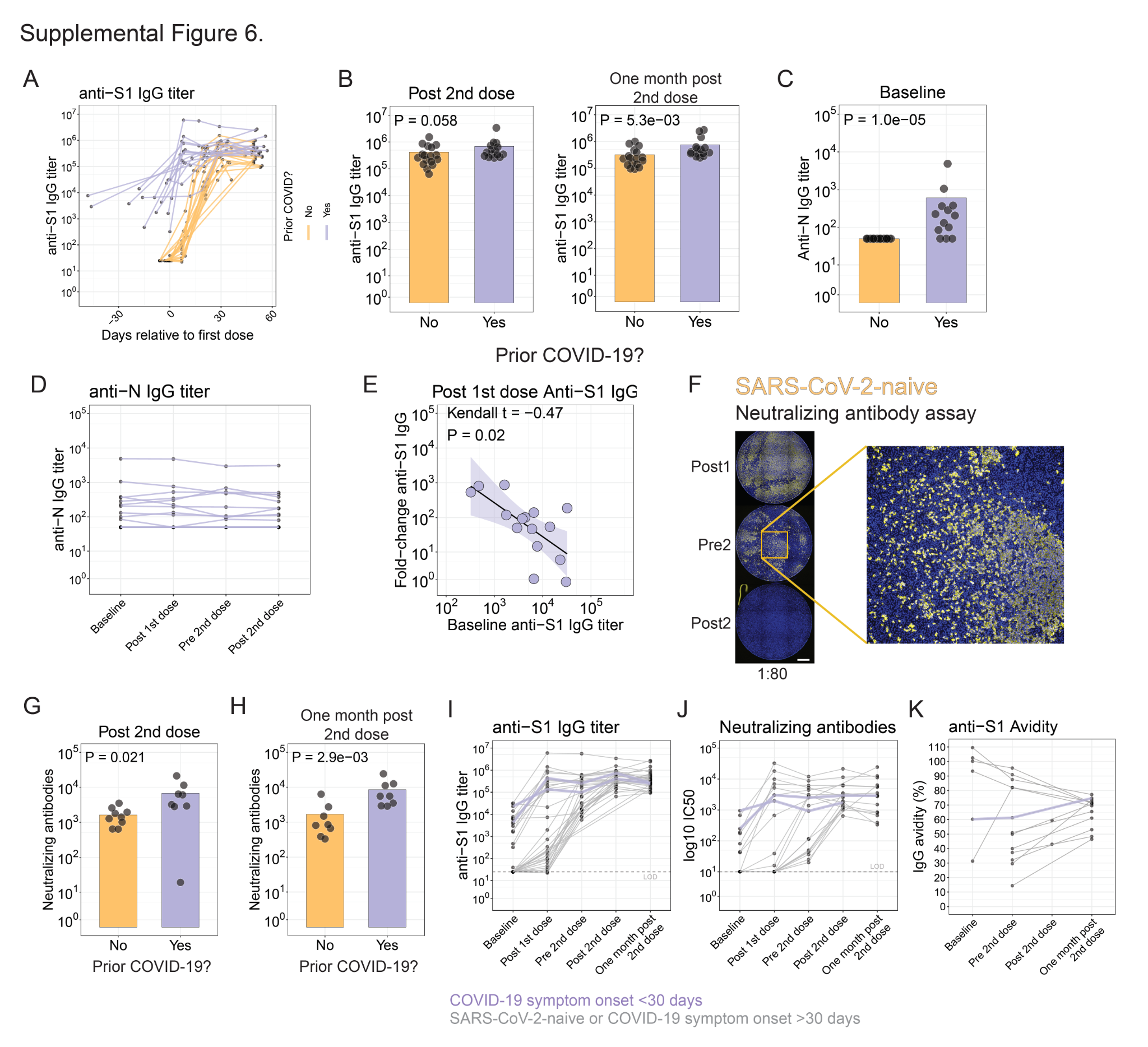

#### Fig. S6. SARS-CoV-2-experienced individuals’ robust anti-S1 binding and neutralizing antibodies responses after vaccination.

(**A**) Anti-S1 IgG serum antibody titers over time measured in days relative to the first vaccination. (**B**) Anti-S1 IgG titer at Post 2nd dose (left) or at the One month post 2nd dose time point (right). (**C**) Anti-nucleocapsid IgG serum antibody titers. (**D**) Correlation between fold-change in anti-S1 IgG serum antibody titers, assessed as one week after vaccination compared to baseline, compared to the baseline anti-S1 IgG serum antibody titers, for SARS-CoV-2-experienced adults. (**E**) Fold-change in antibody titers in SARS-CoV-2-experienced adults post 1st dose relative to titers at baseline. (**F**) Example of neutralizing antibody assay shown for the same SARS-CoV-2-naive participant longitudinally at 1:80 dilution (left), with magnification of well image (right). Scale bar indicates 100 um. (**G**) Neutralizing antibody titers post 2nd dose (*P*=0.02; Wilcoxon test). (**H**) Neutralizing antibody titers one month after the 2nd dose (*P*=2.9x10^-3^; Wilcoxon test). (**I to K**) Summary graphs of the anti-S1 IgG antibody titers (I), plasma neutralizing antibody titers (J), anti-S1 IgG antibody avidity (K) for both cohorts highlighting the participants with recent COVID diagnostic in blue.

#### Table S1. Participant demographics.

|  | **SARS-CoV-2-naive** | **SARS-CoV-2-experienced** |
| --- | --- | --- |
| **Number of participants** | 21 | 15 |
| **Age** |  |  |
| **Median** | 39 | 43 |
| **Range** | 21 – 65 | 24 – 60 |
| **Sex (% Female)** | 48% | 67% |
| **Race (%)** |  |  |
| **White or Caucasian** | 76% | 87% |
| **Asian** | 19% | 13% |
| **Black or African-American** | 5 % | 0 % |
| **Days between diagnosis of COVID-19 and first dose of vaccine** |  |  |
| **Median** |  | 282 |
| **Range** |  | 21 – 359 |

#### Table S2. Clinical COVID-19 disease among SARS-CoV-2-experienced participants.

| **Participant** | **Number of days between onset of COVID-19 symptoms and first dose of**  **Vaccine** | **COVID-19 diagnostic test** | **WHO COVID-19 Severity Score** *(71)* | **Days Hospitalized** | **Outcome** |
| --- | --- | --- | --- | --- | --- |
| CV-043 | 360 | NAAT (PCR) | 7 | 17 | Required intubation, treated with immunomodulator,  improved and was discharged from hospital |
| CV-025 | 358 | anti-S1 IgG ELISA | 1 | 0 | Asymptomatic |
| CV-039 | 323 | NAAT (PCR) | 2 | 0 | Convalescent |
| CV-010 | 301 | Commercial antibody test | 2 | 0 | Convalescent |
| CV-028 | 291 | anti-S1 IgG ELISA | 2 | 0 | Convalescent |
| CV-034 | 290 | NAAT (PCR) | 2 | 0 | Convalescent |
| CV-015 | 285 | NAAT (PCR) | 2 | 0 | Convalescent |
| CV-005 | 284 | Commercial antibody test | 2 | 0 | Convalescent |
| CV-020 | 282 | NAAT (PCR) | 2 | 0 | Convalescent |
| CV-027 | 281 | NAAT (PCR) | 2 | 0 | Convalescent |
| CV-026 | 279 | NAAT (PCR) | 2 | 0 | Convalescent |
| CV-033 | 275 | anti-S1 IgG ELISA | 1 | 0 | Asymptomatic |
| CV-014 | 267 | NAAT (PCR) | 2 | 0 | Convalescent |
| CV-018 | 28 | NAAT (PCR) | 2 | 0 | Convalescent |
| CV-016 | 21 | NAAT (PCR) | 2 | 0 | Convalescent |

#### Table S3. Antibodies used for flow cytometry experiments.

| **Target** | **Fluorochrome** | **Clone** | **Manufacturer** | **Catalog #** |
| --- | --- | --- | --- | --- |
| **Live/Dead Blue** | - | - | Invitrogen | L23105 |
| **CD3** | APC/Fire 810 | SK7 | Biolegend | 344857 |
| **CD4** | SparkBlue 550 | SK3 | Biolegend | 344656 |
| **CD4** | SparkViolet 538 | SK3 | Biolegend | 344674 |
| **CD8** | PE-Fire 640 | SK1 | Biolegend | 344761 |
| **CD8** | SparkBlue 550 | SK1 | Biolegend | 344759 |
| **CD11c** | PerCP | Bu15 | Biolegend | 337234 |
| **CD14** | BUV805 | M5E2 | BD | 612902 |
| **CD16** | BV480 | 3G8 | BD | 566171 |
| **CD19** | BUV496 | SJ25C1 | BD | 612939 |
| **CD20** | APC | 2H7 | Biolegend | 302309 |
| **CD21** | PE-Cy5 | B-ly4 | BD | 551064 |
| **CD23** | BUV615 | M-L233 | BD | 751104 |
| **CD24** | PE-Cy7 | ML5 | Biolegend | 311120 |
| **CD25** | BUV563 | 2A3 | BD | 612919 |
| **CD27** | SB702 | O323 | Invitrogen | 67-0279-42 |
| **CD38** | Qdot655 | HIT2 | Invitrogen | Q22150 |
| **CD40** | BV510 | 5C3 | Biolegend | 334330 |
| **CD45RA** | Spark NIR 685 | HI100 | Biolegend | 304168 |
| **CD56** | BV570 | 5.1H11 | Biolegend | 362539 |
| **CD69** | PE-Dazzle | FN50 | Biolegend | 310942 |
| **CD71** | SB780 | OKT9 | Invitrogen | 78-0719-42 |
| **CD123** | BV650 | 7G3 | BD | 563405 |
| **CD134 (OX40)** | BV421 | ACT35 | Biolegend | 350014 |
| **CD137 (41BB)** | PE | 4B4-1 | Biolegend | 309803 |
| **CD137** | BV750 | 4B4-1 | BD | 747353 |
| **CD138** | PacBlue | MI15 | Biolegend | 356531 |
| **CD150 (CTLA4)** | BV421 | BNI3 | Biolegend | 369606 |
| **CD183 (CXCR3)** | BV750 | 1C6 | BD | 746895 |
| **CD185 (CXCR5)** | BB515 | RF8B2 | BD | 564624 |
| **CD197 (CCR7)** | BV605 | G043H7 | Biolegend | 353224 |
| **CD200 (OX2)** | PE-Cy7 | OX-104 | Biolegend | 329211 |
| **CD278 (ICOS)** | APC-Fire750 | C398.4A | Biolegend | 313536 |
| **CD279 (PD-1)** | BB700 | EH12.1 | BD | 566460 |
| **HLA-DR** | BUV661 | G46-6 | BD | 612980 |
| **IgM** | BUV395 | G20-127 | BD | 563903 |
| **IgD** | BUV737 | IA6-2 | BD | 612798 |
| **Foxp3** | PE-Cy5.5 | PCH101 | Invitrogen | 35-4776-42 |
| **Tbet** | PE-Cy7 | 4B10 | Biolegend | 644823 |
| **Eomes** | PE-eF610 | WD1928 | Invitrogen | 61-4877-42 |
| **GzmB** | A700 | GB11 | BD | 561016 |
| **Ki67** | BUV395 | B56 | BD | 564071 |
| **IgG** | PerCP-Vio700 | IS11-3B2.2.3 | Miltenyi | 130-119-880 |
| **TNF** | BUV396 | MAb11 | BD | 563996 |
| **IFNγ** | BV480 | B27 | BD | 566176 |
